# Metabolomic ageing across mental and behavioural disorders

**DOI:** 10.1101/2025.06.19.25329938

**Authors:** Julian Mutz, Lachlan Gilchrist, Andrea G. Allegrini, Sandra Sanchez Roige, Cathryn M Lewis

## Abstract

**Background:** Individuals with mental and behavioural disorders face increased risk of age-related diseases and premature mortality. Accelerated biological ageing may contribute to this disparity. We investigated differences in metabolomic ageing between individuals with and without mental disorders.

**Methods:** The UK Biobank is a community-based health study of middle-aged and older adults. Mental disorders were identified from hospital inpatient, primary care, death registry and self-reported physician diagnosis data. Plasma metabolites were profiled using the Nightingale Health platform. We examined differences in MileAge delta, the difference between metabolite-predicted and chronological age, across broad ICD-10 diagnostic groups and for 45 individual diagnoses. We further investigated sex-specific associations and tested whether polygenic scores for mental disorders were associated with MileAge delta.

**Results:** Amongst 225,212 participants (54% female; mean age = 56.97 years), 38,524 had at least one mental disorder diagnosis preceding baseline. Substance use, psychotic, affective and neurotic disorders were associated with a metabolite-predicted age exceeding chronological age. In contrast, obsessive-compulsive and eating disorders were associated with a younger MileAge, particularly in females. Associations were generally stronger in males, with several diagnoses showing sex-specific patterns. Higher genetic liability to major depression, autism and ADHD was associated with a MileAge exceeding chronological age, whereas psychosis, tobacco use disorder, obsessive-compulsive disorder and anorexia nervosa polygenic scores were associated with a younger MileAge.

**Conclusions:** Metabolomic ageing varies across mental disorders, with direction and strength of association differing by diagnosis and sex. These findings highlight the heterogeneity of biological ageing across mental disorders and contribute to our understanding of the biological processes linking mental disorders to excess morbidity and premature mortality.

## Introduction

Population ageing is a global health challenge, linked to rising chronic disease burden, disability and healthcare costs (Partridge, Deelen, & Slagboom, 2018). Individuals with mental disorders face increased risks of age-related diseases and reduced life expectancy compared to the general population (Plana-Ripoll et al., 2019). Besides elevated suicide and fatal accident rates (Chan, Tong, Wong, Chen, & Chang, 2022; Khan, Faucett, Morrison, & Brown, 2013) and unhealthy behaviours such as smoking (Taylor et al., 2023), these disparities may reflect differences in the pace of biological ageing (Wolkowitz, 2018).

While chronological age increases uniformly, individuals vary in the rate at which age-related molecular and cellular damage accumulate, impairing function and increasing disease susceptibility. Mental disorders are associated with age-related functional, molecular and clinical markers, including lower grip strength (Ashdown-Franks et al., 2019; Mutz, Hoppen, Fabbri, & Lewis, 2022; Mutz & Lewis, 2021; Mutz, Young, & Lewis, 2022), shorter telomeres (Darrow et al., 2016; Mutz & Lewis, 2022), elevated inflammation (Pitharouli et al., 2021) and increased frailty (Mutz, Choudhury, Zhao, & Dregan, 2022; Pearson et al., 2022).

Advances in computing and the availability of population-scale molecular data have enabled the development of ageing clocks (Horvath & Raj, 2018; Rutledge, Oh, & Wyss-Coray, 2022). Ageing clocks capture the relationship between biological data (e.g., plasma metabolites) and a dependent variable (typically chronological age or mortality) and are used to predict a person’s age. The difference between predicted age and chronological age provides a proxy of biological ageing.

Metabolomics, the profiling of small molecules produced by cellular processes, has become a powerful tool in biomedical research (Würtz et al., 2017). Metabolite levels provide insights into an organism’s physiological state, reflecting internal and external exposures, including genetic, lifestyle and environmental factors (Kettunen et al., 2016; Shin et al., 2014). The pathophysiology of many age-related diseases, including those more prevalent amongst individuals with mental disorders, involves changes in the metabolome (Pietzner et al., 2021). Metabolomics may help decipher the molecular mechanisms of ageing (Panyard, Yu, & Snyder, 2022) and elucidate how these processes may be altered in mental disorders.

Research at the intersection of metabolomics, biological ageing and psychiatry recently emerged, with most studies focused on mood and stress or anxiety-related disorders. Robinson et al. (2020) showed that depression, anxiety, post-traumatic stress disorder and heavy alcohol use were associated with accelerated metabolomic ageing in police officers (*N* = 2212; 60.5% male) (Robinson et al., 2020). However, neither the presence nor severity of depression was associated with metabolomic ageing in a Dutch cohort (*N* = 2910; 66% female) (Jansen et al., 2021), suggesting that findings may differ by population or methodology. Other mental disorders remain underexplored at scale.

The UK Biobank holds one of the largest metabolomics datasets globally. Together with its extensive health record linkage, it represents a key resource to investigate associations between ageing and mental disorders. We developed MileAge, a metabolomic ageing clock trained on plasma metabolite data from over 225,000 individuals. Individuals whose metabolite-predicted age exceeded their chronological age had a higher risk of mortality, greater likelihood of frailty, shorter telomeres and an increased risk of dementia (Mutz, Gilchrist, Pain, Proitsi, & Lewis, 2025; Mutz, Iniesta, & Lewis, 2024).

Here, we examined whether individuals with mental or behavioural disorders show differences in metabolomic ageing. Specifically, we sought to: (1) quantify differences in MileAge delta—the difference between metabolite-predicted and chronological age— between individuals with and without mental disorders; (2) identify possible sex-specific differences; and (3) investigate whether genetic vulnerability to mental disorders contributes to differences in metabolomic ageing. A further objective was to compare associations across broad diagnostic groups and individual diagnoses to determine whether patterns of metabolomic ageing were unique to certain mental disorders or reflected transdiagnostic processes.

## Methods

### Study population

The UK Biobank (UKB) is a community-based health study of over 500,000 individuals aged 37–73 at baseline (Bycroft et al., 2018). Individuals registered with the UK National Health Service and living near one of 22 assessment centres were invited to participate. Between 2006 and 2010, participants reported on sociodemographic factors, health behaviours and medical history, underwent physical examination and provided blood and urine samples. Hospital inpatient records are available for most participants, and primary care records are available for approximately 230,000 participants.

### Mental and behavioural disorders

Mental and behavioural disorders were identified using UKB data fields 130836–130843 (showcase category 1712), which integrate primary care, hospital inpatient, death registry and self-reported physician diagnosis data. International Classification of Diseases (10th revision; ICD-10) codes were extracted from hospital and death registry data. ICD-9 codes from hospital records and Read v2 or CTV3 codes from primary care were mapped to ICD-10. Self-reported physician diagnoses from baseline were also mapped to ICD-10, with occurrence dates interpolated as mid-year. Only diagnoses with a first occurrence date prior to baseline were included. We considered the following ICD-10 codes: F10–F19 (mental and behavioural disorders due to psychoactive substance use), F20–F29 (schizophrenia, schizotypal and delusional disorders), F30–F39 (mood [affective] disorders), F40–F48 (neurotic, stress-related and somatoform disorders), F50–F59 (behavioural syndromes associated with physiological disturbances and physical factors), F60–F69 (disorders of adult personality and behaviour), F70–F79 (mental retardation), F80–F89 (disorders of psychological development), F90–F98 (behavioural and emotional disorders with onset usually occurring in childhood and adolescence) and F99 (unspecified mental disorder). Organic mental disorders (F00–F09) were excluded due to their distinct nature and clinical profile. Diagnoses were not mutually exclusive; individuals with multiple diagnoses were included in all relevant analyses. A binary ‘any disorder’ phenotype was also derived, defined as having at least one ICD-10 code within F10–F99. The comparison group comprised individuals without any recorded mental disorder.

### Metabolomic ageing (MileAge) clock

Metabolomic biomarkers were quantified using nuclear magnetic resonance (NMR) spectroscopy on non-fasting plasma samples collected at baseline. The Nightingale Health platform ascertains 168 circulating metabolites in absolute concentration units using a standardised high-throughput protocol (Würtz et al., 2017). Technical variation was removed using the ‘ukbnmr’ R package (algorithm v2) (Ritchie et al., 2023). In a prior study (Mutz et al., 2024), we developed a metabolomic clock using a Cubist rule-based regression model. Individual-level age predictions were aggregated from the ten test sets of the outer loop of the nested cross-validation to avoid overfitting. Metabolomic age (MileAge) delta represents the difference between metabolite-predicted and chronological age, with positive values indicating an older biological ageing profile (Mutz et al., 2024).

### Polygenic scores

Genotype quality control steps are described in Supplementary Methods. Polygenic scores for mental disorders were calculated using the GenoPred pipeline (Pain, Al-Chalabi, & Lewis, 2024). Each score aligned broadly with the ICD-10 codes reported above. Genome-wide association study (GWAS) summary statistics were obtained from FinnGen, the Psychiatric Genomics Consortium (PGC) and other sources (Table S1), predominantly from individuals of European ancestry. Linkage disequilibrium was estimated using a combined 1000 Genomes Project phase 3 (Auton et al., 2015) and Human Genome Diversity Project (Bergström et al., 2020) reference panel, restricted to 1,204,449 HapMap3 variants. MegaPRS implemented in LDAK v5.1 was used for polygenic scoring, applying the BLD-LDAK heritability model, which incorporates SNP weighting based on allele frequency, linkage disequilibrium and functional annotations to improve predictive performance (Zhang, Privé, Vilhjálmsson, & Speed, 2021). The best-performing polygenic score for each disorder phenotype was selected via pseudo-validation to optimise hyperparameters without requiring individual-level data. Scores were calculated within assigned ancestry groups (matching individuals to five populations in the *N* = 3313 reference data with a probability ≥ 0.95) and scaled to units of standard deviation from each reference population mean (Pain et al., 2021).

### Covariates

A directed acyclic graph (DAG) was used to visualise relationships between mental disorders (exposure), MileAge delta (outcome) and potential confounders: chronological age, sex, highest educational/professional qualification, gross annual household income, ethnicity, cohabitation with spouse/partner, neighbourhood deprivation (Townsend deprivation index) and fasting time (Figure S1). Although genetic confounding was not included in the DAG, we performed separate analyses to evaluate associations between polygenic scores for mental disorders and MileAge delta. These analyses were additionally adjusted for genotype batch number, assessment centre and the first six genetic principal components (derived within UKB) to account for population stratification (Coleman et al., 2020).

### Exclusion criteria

We excluded females who were pregnant or unsure about pregnancy status (due to gestational changes in metabolite levels), individuals with mismatched genetic and self-reported sex, those with missing or outlier metabolite values (defined as values 4× the interquartile range from the median). Individuals with a first occurrence date after the baseline assessment, with no event date or with implausible dates (prior to, matching or in the same calendar year as their birth date or beyond the follow-up period) were also excluded.

### Statistical analyses

Analyses were performed in R (version 4.3.0). Descriptive statistics included means and standard deviations or counts and percentages. Differences in MileAge delta between individuals with mental disorders and the comparison group were estimated using ordinary least squares regression (MileAge delta ∼ diagnosis + covariates). We fitted a minimally adjusted model that included chronological age and sex (Model 1) and a fully adjusted model that also included education, income, ethnicity, cohabitation, deprivation and fasting time (Model 2). MileAge delta was the dependent variable in all models. We first examined broad diagnostic groups (F10–F99, and subgroups F10–F19 through F99), followed by 45 individual diagnoses with a minimum sample size of ten individuals. *P*-values were adjusted for multiple testing using the Benjamini-Hochberg method, with a two-tailed test and a false discovery rate of 5%. To explore sex differences, we performed similar analyses in males and females separately. Associations between polygenic scores for mental disorders and MileAge delta were estimated using ordinary least squares regression (MileAge delta ∼ polygenic score + covariates). These analyses were performed across the full sample and within ancestry groups.

### Sensitivity analyses

To assess the robustness of phenotypic associations, we excluded individuals with mental disorders identified only from self-reported physician diagnosis data due to potential recall bias. Given that only 45% of the sample had linked primary care data, we also performed analyses within this subset to reduce misclassification bias.

## Results

### Sample characteristics

Of 274,315 participants with metabolomic data, 247,861 had complete data (Figure S1). After excluding individuals with possible pregnancy, discordant genetic and self-reported sex or outlier metabolite values, the final sample comprised 225,212 participants (54% female; mean age = 56.97 years, SD = 8.10) (Table 1). Of these, 38,524 (17.1%) had at least one mental disorder diagnosis; the remaining 186,688 (82.9%) served as the comparison group. The most common diagnostic categories were substance use (5.0%, *N* = 11,161), affective (9.0%, *N* = 20,360) and neurotic disorders (6.2%, *N* = 13,916).

**Table 1.** Sample characteristics.

|  | Analytical sample |  |  |
| --- | --- | --- | --- |
| | No diagnosis<br>($N=186,688$) | Any diagnosis<br>($N=38,524$) | Full sample<br>($N=225,212$) |
| Age, mean (SD) | 57.08 (8.13) | 56.45 (7.95) | 56.97 (8.10) |
| Sex |  |  |  |
| Female | 98456 (52.7%) | 23075 (59.9%) | 121531 (54.0%) |
| Male | 88232 (47.3%) | 15449 (40.1%) | 103681 (46.0%) |
| Ethnicity |  |  |  |
| White | 176439 (94.5%) | 36981 (96.0%) | 213420 (94.8%) |
| Mixed | 1017 (0.5%) | 234 (0.6%) | 1251 (0.6%) |
| Black | 2841 (1.5%) | 339 (0.9%) | 3180 (1.4%) |
| Asian | 3371 (1.8%) | 496 (1.3%) | 3867 (1.7%) |
| Chinese | 592 (0.3%) | 48 (0.1%) | 640 (0.3%) |
| Other | 1601 (0.9%) | 255 (0.7%) | 1856 (0.8%) |
| Missing<sup>1</sup> | 827 (0.4%) | 171 (0.4%) | 998 (0.4%) |
| Highest qualification |  |  |  |
| None | 30884 (16.5%) | 7851 (20.4%) | 38735 (17.2%) |
| O levels/GCSEs/CSEs | 50076 (26.8%) | 10624 (27.6%) | 60700 (27.0%) |
| A levels/NVQ/HND/HNC<sup>2</sup> | 42756 (22.9%) | 8768 (22.8%) | 51524 (22.9%) |
| Degree | 60859 (32.6%) | 10819 (28.1%) | 71678 (31.8%) |
| Missing<sup>1</sup> | 2113 (1.1%) | 462 (1.2%) | 2575 (1.1%) |
| Household income<sup>3</sup> |  |  |  |
| Very low | 33380 (17.9%) | 10499 (27.3%) | 43879 (19.5%) |
| Low | 40867 (21.9%) | 8803 (22.9%) | 49670 (22.1%) |
| Medium | 42571 (22.8%) | 7832 (20.3%) | 50403 (22.4%) |
| High | 33854 (18.1%) | 4835 (12.6%) | 38689 (17.2%) |
| Very high | 8833 (4.7%) | 952 (2.5%) | 9785 (4.3%) |
| Missing<sup>1</sup> | 27183 (14.6%) | 5603 (14.5%) | 32786 (14.6%) |
| Cohabitation |  |  |  |
| With partner | 139874 (74.9%) | 24765 (64.3%) | 164639 (73.1%) |
| Single | 14242 (7.6%) | 4108 (10.7%) | 18350 (8.1%) |
| Missing<sup>1</sup> | 32572 (17.4%) | 9651 (25.1%) | 42223 (18.7%) |
| Townsend deprivation |  |  |  |
| Q1 | 39991 (21.4%) | 6735 (17.5%) | 46726 (20.7%) |
| Q2 | 39381 (21.1%) | 7131 (18.5%) | 46512 (20.7%) |
| Q3 | 37660 (20.2%) | 7508 (19.5%) | 45168 (20.1%) |
| Q4 | 36186 (19.4%) | 7807 (20.3%) | 43993 (19.5%) |
| Q5 | 33261 (17.8%) | 9278 (24.1%) | 42539 (18.9%) |
| Missing<sup>1</sup> | 209 (0.1%) | 65 (0.2%) | 274 (0.1%) |
| Fasting time |  |  |  |
| Less than 8 hours | 179744 (96.3%) | 36686 (95.2%) | 216430 (96.1%) |
| At least 8 hours | 6940 (3.7%) | 1838 (4.8%) | 8778 (3.9%) |
| Missing<sup>1</sup> | 4 (0.0%) | 0 (0.0%) | 4 (0.0%) |

*Note:* Numbers shown are counts and percentages unless indicated otherwise. SD = standard deviation. GCSEs = general certificate of secondary education; CSE = certificate of secondary education; NVQ = national vocational qualification; HND = higher national diploma; HNC = higher national certificate. <sup>1</sup> Missing data may include “do not know” or “prefer not to answer”.
<sup>2</sup> Also includes other professional qualifications. <sup>3</sup> Annual household income groups (defined at baseline): very low (<£18,000), low (£18,000–£30,999), middle (£31,000–£51,999), high (£52,000–£100,000) and very high (>£100,000).

### Age of onset relative to baseline

Age of onset relative to baseline distributions are shown in Figure 1 and S3. Median time from onset to baseline ranged from 6.77 years prior (IQR = 10.28) for substance use disorders to 18.95 years prior (IQR = 27.90) for child and adolescent behavioural/emotional disorders (Table S2). Data for individual diagnoses are provided in Table S3 and Figures S4-S11.

**Figure 1.**
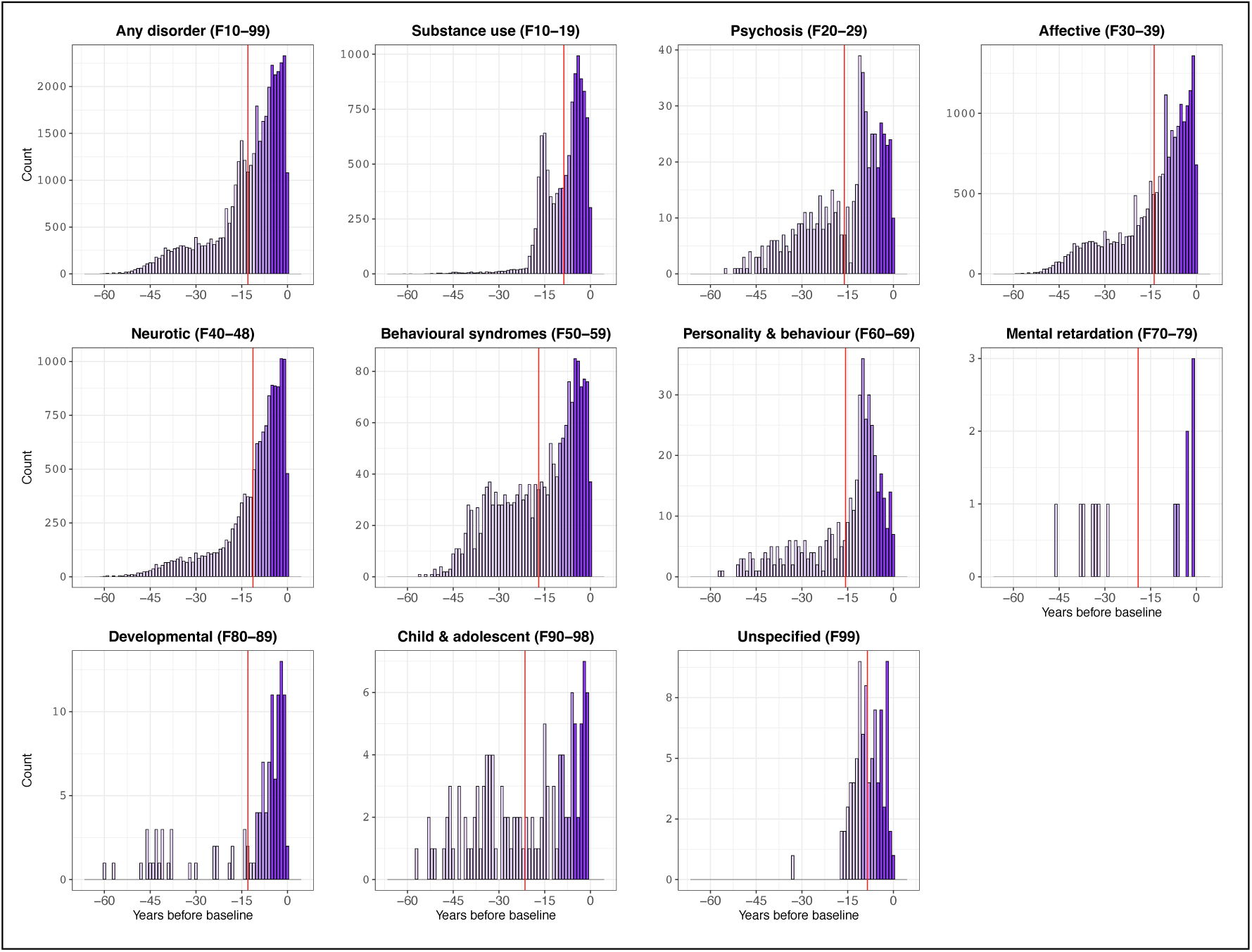
Age of onset differences. Differences in age of onset of mental/behavioural disorders from the baseline assessment date, for groups of two-digit ICD-10 codes. ICD-10 = International Classification of Diseases, 10th Revision. Vertical red lines show the mean difference in age of onset.

### Mental/behavioural disorders and MileAge delta

Across broad diagnostic groups, individuals with substance use, psychotic, affective or neurotic disorders had a metabolite-predicted age (MileAge) exceeding their chronological age (Figure 2A). In age and sex-adjusted models, associations with MileAge delta ranged from *β* = 0.152 (95% CI 0.088–0.217, *p* < 0.001) for neurotic disorders to *β* = 0.740 (95% CI 0.436–1.045, *p* < 0.001) for psychosis. Conversely, behavioural syndromes were associated with a younger metabolite-predicted age than chronological age (*β* = -0.210, 95% CI -0.387 to -0.034, *p* = 0.036). All associations remained statistically significant after full adjustment, though most attenuated (Table 2).

**Figure 2.**
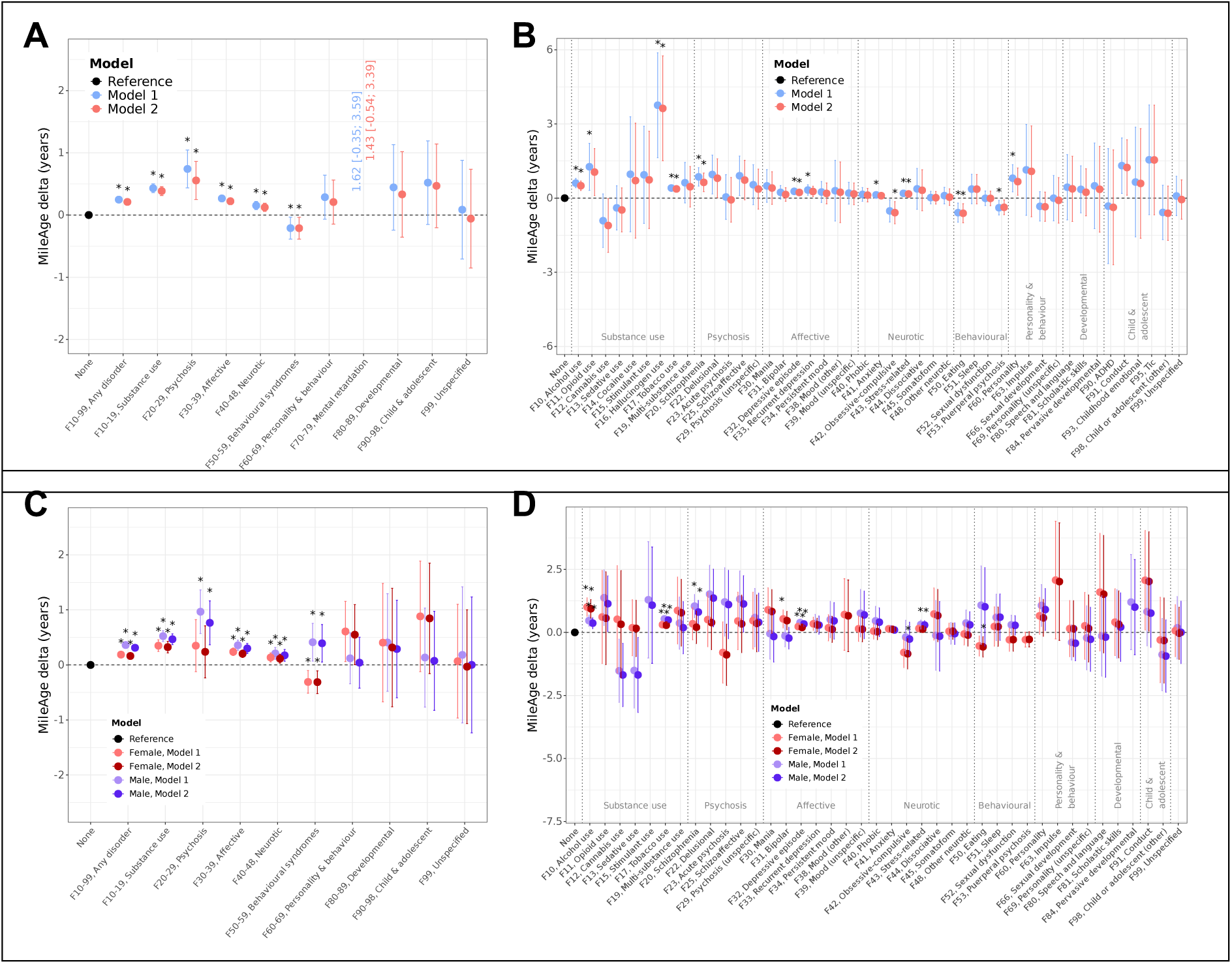
MileAge delta and mental/behavioural disorders. **(A)** Differences in MileAge delta between individuals with and without mental/behavioural disorders, for groups of two-digit ICD-10 codes. **(B)** Differences in MileAge delta between mental/behavioural disorders, for individual two-digit ICD-10 codes. ADHD = attention-deficit/hyperactivity disorder. **(C)** Sex-stratified differences in MileAge delta between mental/behavioural disorders, for groups of two-digit ICD-10 codes. **(D)** Sex-stratified associations between mental/behavioural disorders and MileAge delta, for individual two-digit ICD-10 codes. **(A** to **D)** ICD-10 = International Classification of Diseases, 10th Revision. Betas and 95% confidence intervals were estimated using linear regression models. Reference group: individuals without mental/behavioural disorders. **(A** and **B)** Model 1–adjusted for chronological age and sex; Model 2–adjusted for chronological age, sex, ethnicity, cohabitation with spouse/partner, highest educational/professional qualification, annual gross household income, Townsend deprivation index and fasting time. Asterisks indicate statistical significance after correcting *p*-values for multiple testing using the Benjamini–Hochberg procedure. **(C** and **D)** Model 1–adjusted for chronological age; Model 2–adjusted for chronological age, ethnicity, cohabitation with spouse/partner, highest educational/professional qualification, annual gross household income, Townsend deprivation index and fasting time. Asterisks indicate statistical significance after correcting *p*-values for multiple testing using the Benjamini–Hochberg procedure within each sex.

**Table 2.**
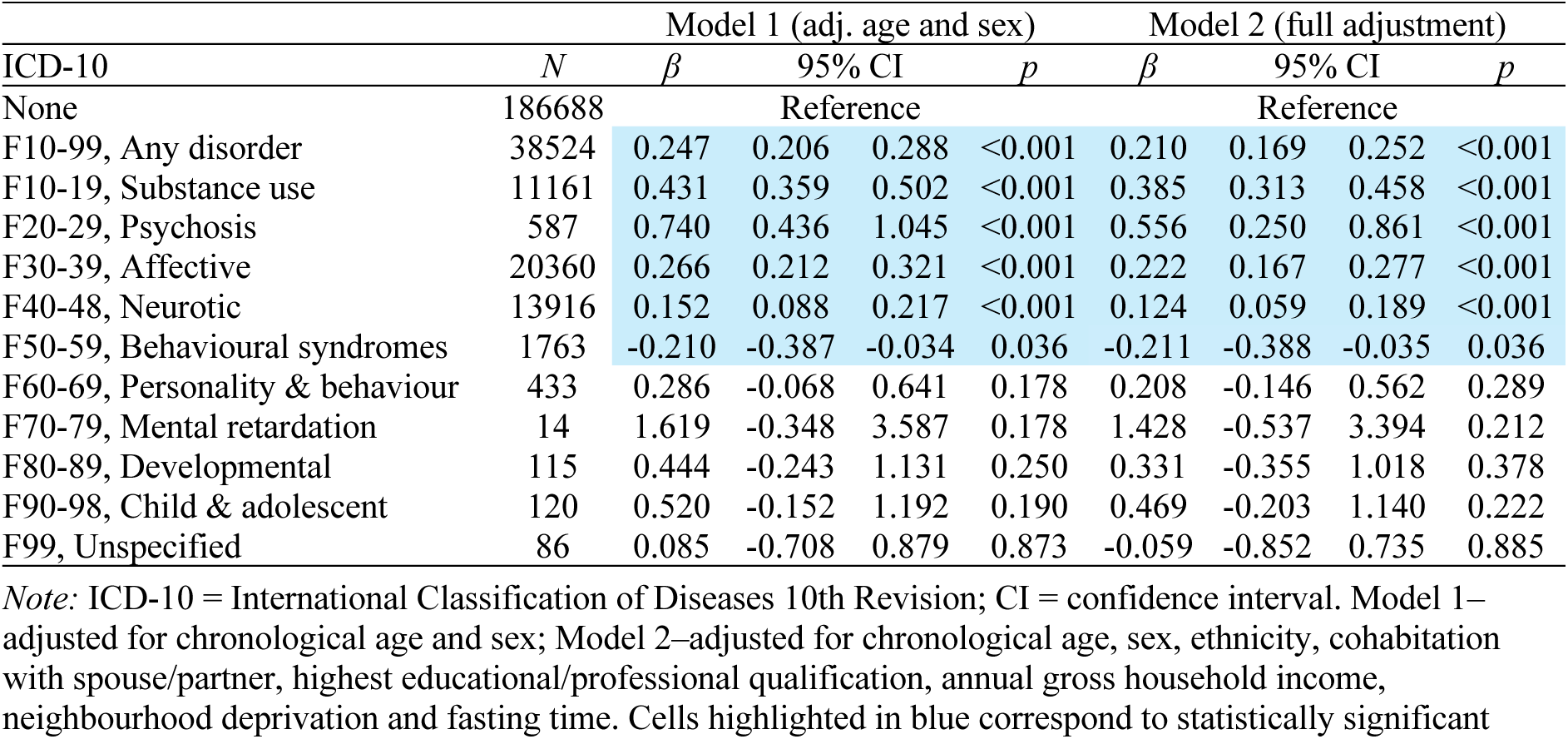
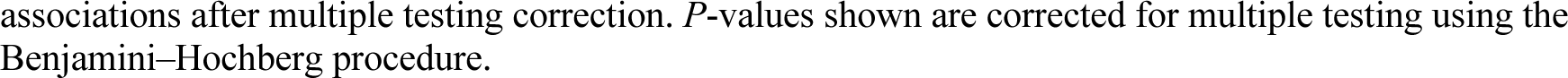
Associations between MileAge delta and mental/behavioural disorders.

Associations with individual diagnoses are shown in Figure 2B. After full adjustment and multiple testing correction, alcohol, hallucinogen and tobacco use disorders, schizophrenia, single-episode depression and stress-related disorders were associated with a metabolite-predicted age exceeding chronological age. Eating disorders were associated with a younger metabolite-predicted age. Full results, including nominal associations, are shown in Table S4.

### Sex-stratified analyses

Associations between broad diagnostic groups and MileAge delta were generally stronger in males (Figure 2C). Behavioural syndromes were associated with a metabolite-predicted age exceeding chronological age in males but with a younger MileAge in females (Table S5). The association between psychosis and metabolomic ageing was not statistically significant in females (*β* = 0.239, 95% CI -0.235–0.712, *p* = 0.431) and weaker than in males (*β* = 0.765, 95% CI 0.368–1.163, *p* < 0.001).

Alcohol and tobacco use disorders were associated with a metabolite-predicted age exceeding chronological age in both sexes (Figure 2D), though effect sizes differed: alcohol use was more strongly associated in females (*β* = 1.008, 95% CI 0.641–1.374, *p* < 0.001) and tobacco use in males (*β* = 0.535, 95% CI 0.429–0.641, *p* < 0.001). Schizophrenia was associated with a MileAge exceeding chronological age in males only (*β* = 0.807, 95% CI 0.345–1.269, *p* = 0.006). Single-episode depression showed stronger associations in males (*β* = 0.332, 95% CI 0.238–0.426, *p* <0.001) than females (*β* = 0.197, 95% CI 0.127–0.268, *p* < 0.001). Associations between stress-related disorders and MileAge delta were statistically significant in males only. Obsessive-compulsive and eating disorders were associated with a MileAge younger than chronological age in females only. Full results are reported in Table S6.

### Polygenic score analysis

We next tested whether genetic liability to mental disorders was associated with metabolomic ageing. Polygenic scores for psychosis were associated with a metabolite-predicted age younger than chronological age (Figure 3A; Table S7). Ancestry-stratified analyses suggested that the association between higher genetic loadings for psychosis and a younger metabolite-predicted age was statistically significant only in Europeans (Figure 3B; Table S8). Polygenic scores for substance use disorders were associated with a MileAge exceeding chronological age in admixed Americans (mixed Indigenous American, European and African ancestry).

**Figure 3.**
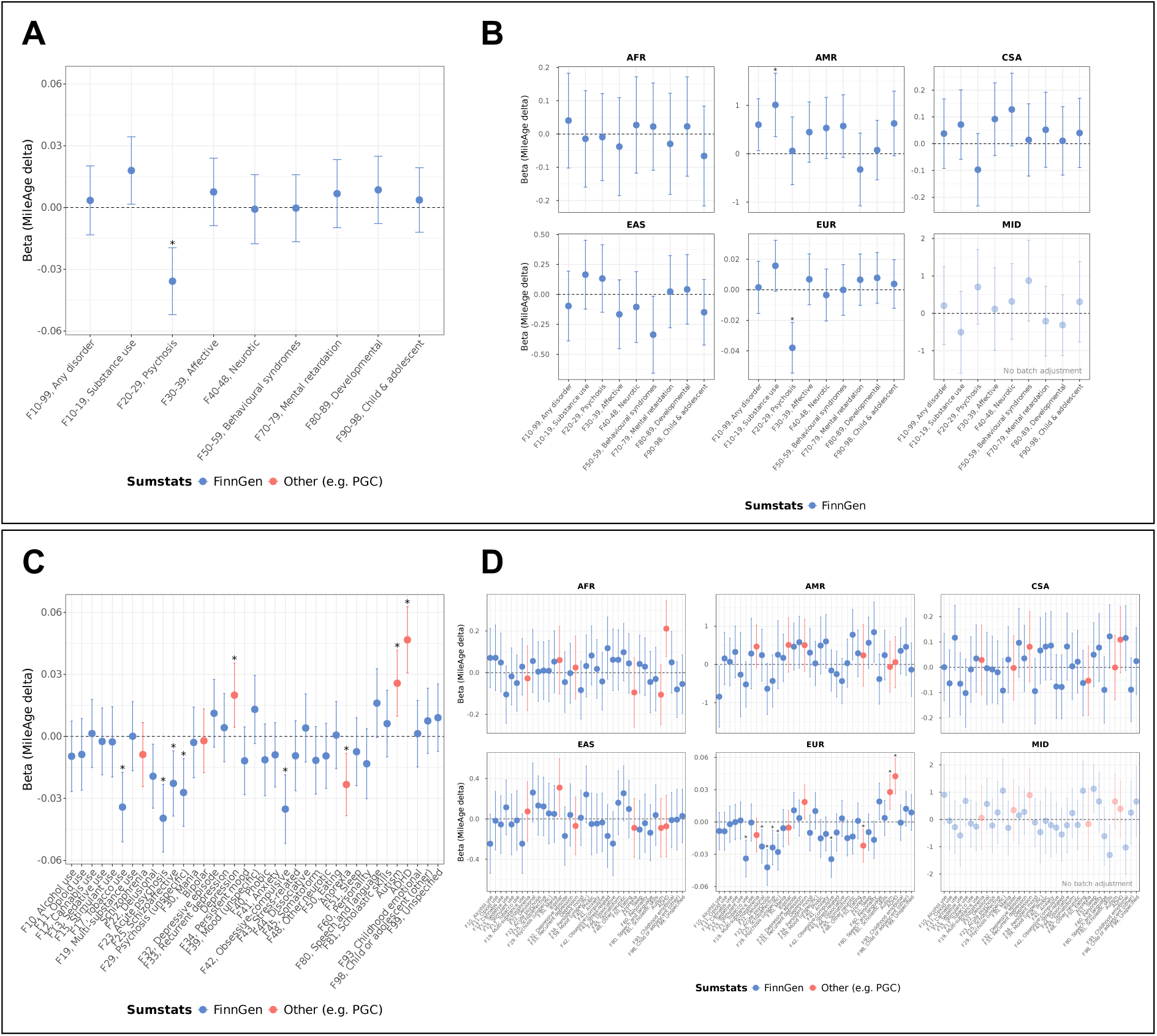
MileAge delta and polygenic scores for mental/behavioural disorders. **(A)** Associations between polygenic scores for groups of mental/behavioural disorders and MileAge delta across ancestries. **(B)** Ancestry-specific associations between polygenic scores for groups of mental/behavioural disorders and MileAge delta. **(C)** Associations between polygenic scores for individual mental/behavioural disorders and MileAge delta across ancestries. **(D)** Ancestry-specific associations between polygenic scores for individual mental/behavioural disorders and MileAge delta. **(A** to **D)** Estimates shown are ordinary least squares regression beta coefficients and 95% confidence intervals. Models were adjusted for chronological age, sex, assessment centre, batch number, the first six genetic principal components and fasting time. Asterisks indicate statistical significance after correcting *p*-values for multiple testing using the Benjamini–Hochberg procedure (separately for groups of diagnoses and individual diagnoses, and separately within each population). PGC = Psychiatric Genomics Consortium. **(A** and **C)** *N* = 219,494. **(B** and **D)** AFR = African; AMR = Admixed American; EAS = East Asian; EUR = European; CSA = Central and South Asian; MID = Middle Eastern. *N* = 3426 (AFR); *N* = 278 (AMR); *N* = 3916 (CSA); *N* = 1012 (EAS); *N* = 210,755 (EUR); *N* = 94 (MID). **(C** and **D)** ADHD = attention-deficit/hyperactivity disorder.

Polygenic scores major depression, autism and attention-deficit/hyperactivity disorder (ADHD) were associated with a MileAge exceeding chronological age (*β* range = 0.020 to 0.047; Table S9). In contrast, higher genetic liability to tobacco use disorder, acute psychosis, schizoaffective disorder, unspecific psychosis, obsessive-compulsive disorder and anorexia nervosa was associated with a MileAge younger than chronological age (*β* range = -0.023 to - 0.040; Figure 3C). After multiple testing correction, these associations were statistically significant only in Europeans (Figure 3D). Nominal associations were observed in other ancestry groups (Table S10). For example, polygenic scores for bipolar disorder (East Asian), other neurotic disorders (admixed American) and ADHD (African) were associated with a metabolite-predicted age exceeding chronological age. Polygenic scores for alcohol use disorder (admixed American), developmental disorders of scholastic skills and childhood emotional disorders (Middle Eastern) were associated with a younger MileAge.

### Sensitivity analysis

Excluding self-reported physician diagnoses (applied to 12 of 45 ICD-10 codes yielded comparable results (Table S11). Sample sizes declined between 15.4% (alcohol use disorder) and 77.2% (bipolar disorder). Some differences emerged: anxiety and obsessive-compulsive disorders became statistically significantly associated with a MileAge exceeding chronological age. Schizophrenia and eating disorders were no longer statistically significantly associated with MileAge delta. For example, the association for schizophrenia declined from *β* =0.638 to *β* = 0.236, with a 61.1% reduction in sample size.

In the subset with primary care data, most estimates were attenuated (Tables S12-S13). Sample sizes reduced for 38 of 45 ICD-10 codes. Some associations, such as for eating disorders, no longer reached statistical significance after full adjustment.

## Discussion

Mental disorders are associated with a greater age-related disease burden and reduced life expectancy. Accelerated biological ageing has been proposed as one mechanism contributing to these disparities, yet evidence linking mental disorders to molecular ageing markers remains limited, particularly at population scale and across diagnostic groups. Using data from over 225,000 individuals, we found that substance use, psychotic, affective and neurotic disorders were associated with a metabolite-predicted age (MileAge) exceeding chronological age. In contrast, behavioural syndromes such as eating disorders were associated with a MileAge younger than chronological age. Sex-stratified analyses identified generally stronger associations in males, with sex differences for in alcohol and tobacco use disorders and behavioural syndromes. In polygenic score analyses, higher genetic liability to major depression, autism and ADHD was associated with a metabolite-predicted age exceeding chronological age, whereas higher genetic liability to tobacco use disorder, psychosis, obsessive-compulsive disorder and anorexia nervosa was associated with a MileAge younger than chronological age. Ancestry-stratified analyses identified that substance use disorder polygenic scores were associated with a metabolite-predicted age exceeding chronological age in individuals of mixed Indigenous American, European and African ancestry.

Most previous studies of biological ageing in mental disorders have focused on mood and stress/anxiety-related disorders, reporting mixed findings across ageing indicators (Han et al., 2021; Liu et al., 2022; Robinson et al., 2020). Few have investigated metabolomic ageing. A UK-based study of police officers found that depression, anxiety and PTSD were associated with accelerated metabolomic ageing (Robinson et al., 2020), which partially aligns with our results. However, a Dutch study reported no association between depression and metabolomic ageing (Jansen et al., 2021). Our observation that depression was associated with altered metabolomic profiles aligns with a Mendelian randomisation study showing that depression may causally affect certain metabolites (Amin et al., 2023). Interestingly, metabolomic profiles were similar in lifetime and recurrent depression, aligning with our observation that associations with MileAge delta were similar for single-episode and recurrent depression. That said, a separate Mendelian randomisation study identified metabolites as potential causal contributors to depression rather than the reverse (Davyson et al., 2023).

Substance use disorders were also associated with a metabolite-predicted age exceeding chronological age, consistent with prior evidence of accelerated epigenetic ageing in alcohol use disorder (Rosen et al., 2018) and higher brain-predicted age amongst those with tobacco and alcohol use (Ning, Zhao, Matloff, Sun, & Toga, 2020). Smoking is known to alter circulating metabolites (Xu et al., 2013), which may partly explain the observation of MileAge exceeding chronological age in tobacco use disorder. Fewer cases were identified for substances other than alcohol and tobacco, highlighting the need for further research.

Interestingly, both obsessive-compulsive and eating disorders were associated with a metabolite-predicted age younger than chronological age in females, consistent with the high genetic correlation between these disorders (Yilmaz et al., 2020). This may reflect a more youthful metabolic profile but could also indicate atypical or dysregulated metabolic states. For example, energy conservation and lipid suppression are common in anorexia nervosa and could bias metabolomic age estimates. Our finding that schizophrenia was associated with a MileAge exceeding chronological age in males aligns with a recent lipidomic study of prefrontal cortex tissue showing age acceleration in schizophrenia and autism (Latumalea, Unfried, Barardo, Gruber, & Kennedy, 2024).

A novel contribution of this study lies in the sex-stratified analyses. Associations were generally stronger in males, although, for instance, females showed stronger associations for alcohol use disorder. Psychosis and stress-related disorders were associated with a metabolite-predicted age exceeding chronological age were only in males, while obsessive-compulsive and eating disorders were associated with a MileAge younger than chronological age only in females. These differences may partly reflect sample size imbalances (e.g., 342 females vs 22 males with eating disorders). They could also indicate sex-specific biological pathways, including hormonal influences or differential health behaviours (e.g., higher smoking rates in men with schizophrenia). Our findings underscore the importance of considering sex in studies of biological ageing in mental disorders.

We also explored associations between metabolomic ageing and genetic liability to mental disorders. Higher polygenic scores for major depression, autism and ADHD were associated with a metabolite-predicted age exceeding chronological age. In contrast, higher polygenic scores for tobacco use disorder, psychosis, obsessive-compulsive disorder and anorexia nervosa were associated with a MileAge younger than chronological age. These associations were modest, corresponding to a maximum difference of ∼2.5 weeks per standard deviation increase in polygenic score. Although current polygenic scores explain only a small proportion of the variance in their target traits, these findings suggest that differences in metabolomic ageing are not driven primarily by common genetic variants linked to these disorders. We also identified diverging results between diagnosis and polygenic scores: for example, while psychosis diagnoses were associated with a MileAge exceeding chronological age, higher polygenic scores for psychosis were associated with a metabolite-predicted age younger than chronological age. These discrepancies may reflect non-genetic influences on biological ageing. While most associations survived multiple testing correction only in individuals of European ancestry, substance use disorder polygenic scores were associated with a metabolite-predicted age exceeding chronological age in admixed Americans. We also identified nominal associations in other populations: for example, bipolar disorder polygenic scores were associated with a MileAge exceeding chronological age in East Asians. Given that our polygenic scores were based on European GWAS summary statistics, these findings should be interpreted cautiously.

Certain limitations must be acknowledged. First, differences in metabolomic ageing may not reflect a causal link between mental disorders and biological age. There is emerging evidence that accelerated biological ageing also predicts incident mental disorders (Jia et al., 2024; Shang et al., 2024; Zeng et al., 2024). However, most participants were 40 to 69 years old at baseline, with age of onset for many diagnoses well preceding the blood sample collection. Of note, duration of illness may impact biological ageing, particularly for chronic conditions such as tobacco use disorder. Future work should model duration of illness to assess potential cumulative effects on biological ageing. Second, although we adjusted for potential confounders, we cannot exclude the possibility of residual confounding. For example, smoking was not explicitly adjusted for because it is on the causal pathway, though it may be partly captured through covariates such as education and deprivation. Unmeasured genetic confounding remains a limitation. Third, some diagnostic misclassification is possible. Individuals with mental disorders were identified mostly from linked health records. Sensitivity analyses largely confirmed our findings, though some associations attenuated and were no longer statistically significant with more restricted case ascertainment. Findings for diagnoses with few cases should be interpreted cautiously. Fourth, given the well documented healthy volunteer bias in the UK Biobank, observed differences in metabolomic ageing represent conservative estimates as the most clinically severe cases are not well represented in the data. Future studies should consider weighting approaches. Finally, most GWAS summary statistics used for polygenic scoring were obtained from the FinnGen study which comprises over 500,000 Finnish participants (Kurki et al., 2023). The portability of these polygenic scores to other ancestry groups is uncertain.

## Conclusion

In conclusion, we identified differences in metabolomic ageing across a broad range of mental and behavioural disorders. Substance use, psychotic, affective and neurotic disorders were associated with a metabolite-predicted age exceeding chronological age. In contrast, obsessive-compulsive disorder and behavioural syndromes such as eating disorders were associated with a MileAge younger than chronological age. Several associations differed by sex: psychosis and stress-related disorders were associated with a metabolite-predicted age exceeding chronological age only in males, while obsessive-compulsive and eating disorders were associated with a MileAge younger than chronological age only in females. These findings highlight the heterogeneity of biological ageing across mental disorders, elucidating our understanding of the biological processes linking mental disorders to excess morbidity and premature mortality.

## Acknowledgments

JM is funded by the King’s Prize Fellowship and a 2024 NARSAD Young Investigator Grant from the Brain & Behavior Research Foundation. LG is funded by the King’s College London DRIVE-Health Centre for Doctoral Training and the Perron Institute for Neurological and Translational Science. CML receives funding through a Wellcome Mental Health Award (226770/Z/22/Z). This work was part-funded by the National Institute for Health and Care Research (NIHR) Maudsley Biomedical Research Centre (BRC). The views expressed are those of the authors and not necessarily those of the NHS, the NIHR or the Department of Health and Social Care. Computational analyses were supported by King’s Computational Research, Engineering and Technology Environment (CREATE). This research has been conducted using data from UK Biobank. Data access permission has been granted under UK Biobank application 45514. We acknowledge the participants and investigators of the FinnGen study for use of their GWAS summary statistics.

## Financial disclosures

CML is a member of the scientific advisory board of Myriad Neuroscience, has received speaker fees from SYNLAB and received consultancy fees from UCB. JM, LG, AGA and SSR declare no conflict of interest.

## Authorship contributions

JM conceived the idea of the study, acquired the data, carried out the analysis, interpreted the findings and wrote the manuscript. LG provided methodological support. LG, AGA, SSR and CML revised the manuscript. All authors read and approved the final manuscript.

## Ethics

Ethical approval for the UK Biobank study has been granted by the National Information Governance Board for Health and Social Care and the NHS North West Multicentre Research Ethics Committee (11/NW/0382). No project-specific ethical approval is needed.

## Data sharing statement

The data used are available to all *bona fide* researchers for health-related research that is in the public interest, subject to an application process and approval criteria. Study materials are publicly available online at http://www.ukbiobank.ac.uk.

## Supplementary material

Supplementary information is available online.

