## Supplementary material for "Metabolomic ageing across mental and behavioural disorders"

This material accompanies the article

12. MileAge delta and individual two-digit ICD-10 codes; excluding self-report ... 29
13. MileAge delta and groups of two-digit ICD-10 codes; primary care linkage .... 30
14. MileAge delta and individual two-digit ICD-10 codes; primary care linkage ... 31

### 1. Supplementary methods

**Imputed genotype data**

Details on genotype calling, quality control and imputation performed centrally by the UK Biobank have been reported elsewhere (Bycroft et al., 2018). Individuals were genotyped using the UK BiLEVE Axiom ($N$ = 49,950) and UK Biobank Axiom ($N$ = 438,427) arrays. The data were imputed using the Haplotype Reference Consortium (HRC) and the combined UK10K and 1000 Genomes Project phase 3 reference panels, with imputed data available for approximately 93,095,623 autosomal SNPs. These were filtered using a minor allele frequency (MAF) threshold of ≥ 1% and INFO score of ≥ 0.4.

**Genotype quality control**

Quality control steps were applied using the GenoPred pipeline (v2.2.11) (Pain, Al-Chalabi, & Lewis, 2024).

GWAS summary statistics: Variants were excluded if they were not identified in the reference dataset, were strand ambiguous, had an INFO score < 0.9 (if available), had out-of-bound $p$-values ($0 < P \leq 1$), had duplicate SNP IDs, had a sample size > 3SD from the median (if available) or had a standard error of zero. MAF thresholds were set at 0.01 for the GWAS summary statistics and the reference dataset, with a MAF difference threshold of 0.2.

Individual-level genotype data: Variants absent from the reference data and duplicate variants were removed. Variants with mismatched RSIDs and variants that were strand-flipped were corrected. Strand-ambiguous variants were excluded. Reference variants absent from the UK Biobank data were inserted as missing to allow reference allele frequency-based imputation during downstream polygenic scoring. Relatedness was estimated using the KING estimator in PLINK v1.9, with unrelated individuals defined using a threshold of $r > 0.044$ (equivalent to removing third-degree relatives and closer).

#### 2. GWAS summary statistics

**Table S1.** Genome-wide association study (GWAS) summary statistics

| Code | N Cases | N Controls | N Total | N Effective | Prop. Cases | Prevalence[1] | PMID |
|---|---|---|---|---|---|---|---|
| Groups of ICD-10 codes | | | | | | | |
| F10–F99 | 138044 | 362304 | 500348 | 399833 | 0.27589598 | 0.27589598 | 36653562 |
| F10–19 | 30806 | 362304 | 393110 | 113568 | 0.07836483 | 0.07836483 | 36653562 |
| F20–29 | 15498 | 362304 | 377802 | 59449 | 0.04102149 | 0.04102149 | 36653562 |
| F30–39 | 65517 | 434831 | 500348 | 227752 | 0.13094286 | 0.13094286 | 36653562 |
| F40–48 | 56552 | 362304 | 418856 | 195666 | 0.13501538 | 0.13501538 | 36653562 |
| F50–59 | 15447 | 484901 | 500348 | 59880 | 0.03087251 | 0.03087251 | 36653562 |
| F60–69 | | | | *N* insufficient | | | |
| F70–79 | 1429 | 362234 | 363663 | 5694 | 0.00392946 | 0.00392946 | 36653562 |
| F80–89 | 5519 | 494829 | 500348 | 21832 | 0.01103032 | 0.01103032 | 36653562 |
| F90–98 | 9640 | 490708 | 500348 | 37817 | 0.01926659 | 0.01926659 | 36653562 |
| F99 | 3449 | 479456 | 482905 | 13697 | 0.00714219 | 0.00714219 | 36653562 |
| Individual ICD-10 codes[2] | | | | | | | |
| F10 | 20597 | 479751 | 500348 | 78996 | 0.04116535 | 0.0283640 | 36653562 |
| F11 | 1444 | 491508 | 492952 | 5759 | 0.00292929 | 0.0038920 | 36653562 |
| F12 | 1168 | 491508 | 492676 | 4661 | 0.00237073 | 0.0052201 | 36653562 |
| F13 | 2668 | 491508 | 494176 | 10614 | 0.00539889 | 0.0019398 | 36653562 |
| F15 | 1077 | 491508 | 492585 | 4299 | 0.00218642 | 0.0008272 | 36653562 |
| F16 | | | | *N* insufficient | | | |
| F17 | 3200 | 495423 | 498623 | 12718 | 0.00641767 | 0.0672100 | 36653562 |
| F19 | 2866 | 491508 | 494374 | 11398 | 0.00579723 | 0.0047489 | 36653562 |
| Schizophrenia | 76755 | 243649 | 320404 | 233471 | 0.23955690 | 0.01 | 35396580 |
| F22 | 2953 | 484776 | 487729 | 11740 | 0.00605459 | 0.0021637 | 36653562 |
| F23 | 5003 | 484776 | 489779 | 19808 | 0.01021481 | 0.0010761 | 36653562 |
| F25 | 3095 | 484776 | 487871 | 12301 | 0.00634389 | 0.0010109 | 36653562 |
| F29 | 8976 | 484776 | 493752 | 35251 | 0.01817917 | 0.01817917 | 36653562 |
| F30 | 1157 | 434831 | 435988 | 4616 | 0.00265374 | 0.0010171 | 36653562 |
| Bipolar disorder | 20352 | 31358 | 51710 | 49367 | 0.39357960 | 0.02 | 31043756 |
| F32 | 59333 | 434831 | 494164 | 208836 | 0.12006743 | 0.1155600 | 36653562 |
| F33 | 26094 | 254393 | 280487 | 94666 | 0.09303105 | 0.0112840 | 36653562 |
| Major depression | 310128 | 1035355 | 1345483 | 829249 | 0.23049570 | 0.12 | 39814019[‡] |
| F34 | 9001 | 434831 | 443832 | 35274 | 0.02028020 | 0.0015478 | 36653562 |
| F38 | | | | *N* insufficient | | | |
| F39 | 3063 | 434831 | 437894 | 12166 | 0.00699484 | 0.00699484 | 36653562 |
| F40 | 6248 | 444414 | 450662 | 24646 | 0.01386405 | 0.0081370 | 36653562 |
| F41 | 35875 | 444414 | 480289 | 132781 | 0.07469461 | 0.0900500 | 36653562 |
| F42 | 2813 | 444414 | 447227 | 11181 | 0.00628987 | 0.0043330 | 36653562 |
| F43 | 18806 | 444414 | 463220 | 72170 | 0.04059842 | 0.0592720 | 36653562 |
| F44 | 1525 | 444414 | 445939 | 6079 | 0.00341975 | 0.0011276 | 36653562 |
| F45 | 7148 | 444414 | 451562 | 28139 | 0.01582950 | 0.0033942 | 36653562 |
| F48 | 1914 | 444414 | 446328 | 7623 | 0.00428833 | 0.0031640 | 36653562 |
| F50 | 3923 | 362304 | 366227 | 15524 | 0.01071194 | 0.0059592 | 36653562 |
| Anorexia nervosa | 16224 | 52460 | 68684 | 49566 | 0.23621220 | 0.01 | 31308545[‡] |
| F51 | 8336 | 362304 | 370640 | 32594 | 0.02249083 | 0.0028453 | 36653562 |
| F52 | | | | *N* insufficient | | | |
| F53 | | | | *N* insufficient | | | |
| F60 | 13045 | 362300 | 375345 | 50367 | 0.03475469 | 0.0058590 | 36653562 |
| F63 | | | | *N* insufficient | | | |
| F66 | | | | *N* insufficient | | | |
| F69 | | | | *N* insufficient | | | |

| | | | | | | | |
|---|---|---|---|---|---|---|---|
| F80 | 2362 | 494829 | 497191 | 9403 | 0.00475069 | 0.0042170 | 36653562 |
| F81 | 1719 | 494829 | 496548 | 6852 | 0.00346190 | 0.0048760 | 36653562 |
| F84 | | | | *N* insufficient | | | |
| Autism | 22458 | 29386 | 51844 | 50918 | 0.43318420 | 0.01 | 32747698 |
| ADHD | 38691 | 186843 | 225534 | 128213 | 0.17155280 | 0.05 | 36702997 |
| F91 | | | | *N* insufficient | | | |
| F93 | 2230 | 490708 | 492938 | 8880 | 0.00452390 | 0.0008979 | 36653562 |
| F95 | | | | *N* insufficient | | | |
| F98 | 2097 | 362304 | 364401 | 8340 | 0.00575465 | 0.00117680 | 36653562 |
| F99 | 3449 | 479456 | 482905 | 13697 | 0.00714219 | 0.00714219 | 36653562 |

*Note:* ADHD = Attention-deficit/hyperactivity disorder; PMID = PubMed reference number. Summary statistics were limited to those with a minimum of 1000 cases. [1] Most prevalence estimates were obtained from https://www.prevalenceuk.com/ and correspond to lifetime prevalence; alternatively, we report estimates reported elsewhere or the proportion of cases (printed in grey). [2] Mapping to two-digits ICD-10 codes may differ for consortia summary statistics (listed without ICD-10 code). [†] Excluding UK Biobank and 23andMe. [‡] Excluding UK Biobank.

3. Directed acyclic graph

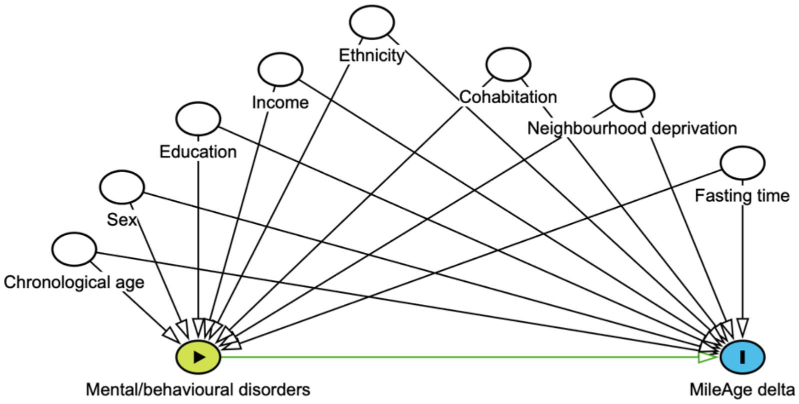

**Figure S1.** Directed acyclic graph (DAG) showing relationships between the exposure (mental/behavioural disorders), outcome (MileAge delta) and select confounders (chronological age, sex, highest educational/professional qualification, gross annual household income, ethnicity, cohabitation with spouse/partner, Townsend deprivation index and fasting time).

#### 4. Study flowchart

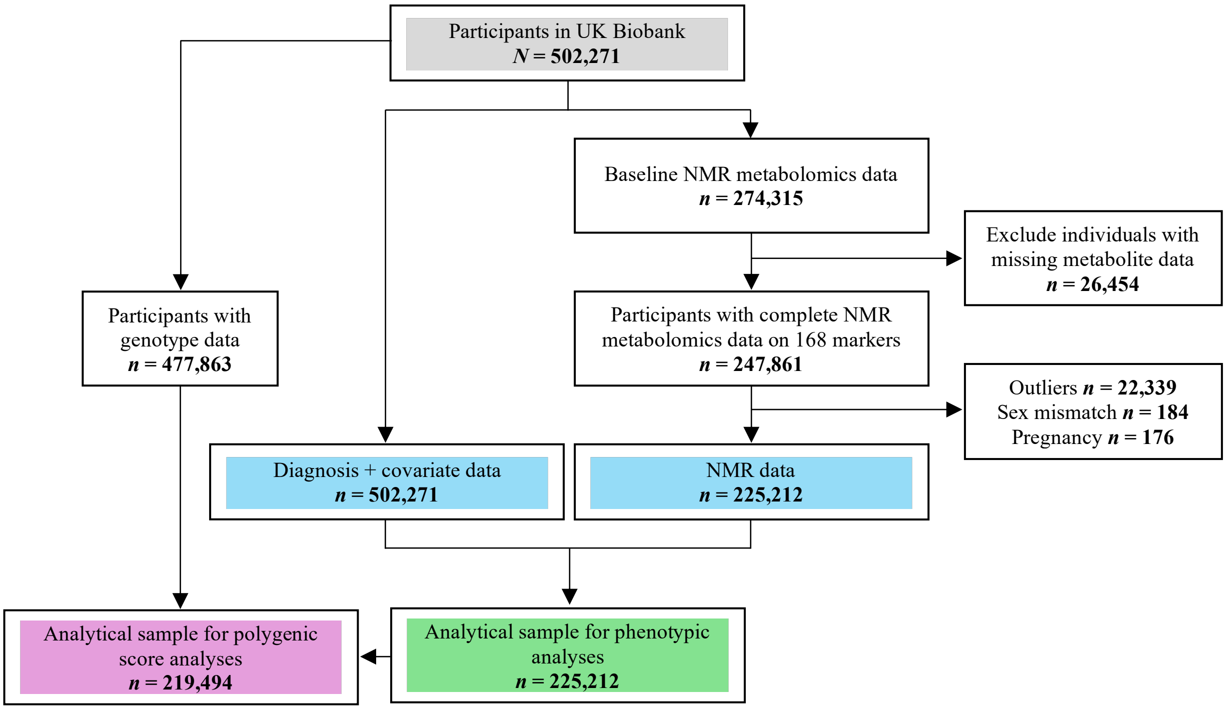

**Figure S2.** Study sample flowchart. Outliers were defined as metabolite values 4×IQR above or below the median. NMR = nuclear magnetic resonance; IQR = interquartile range.

### 5. Age of onset differences for groups of two-digit ICD-10 codes

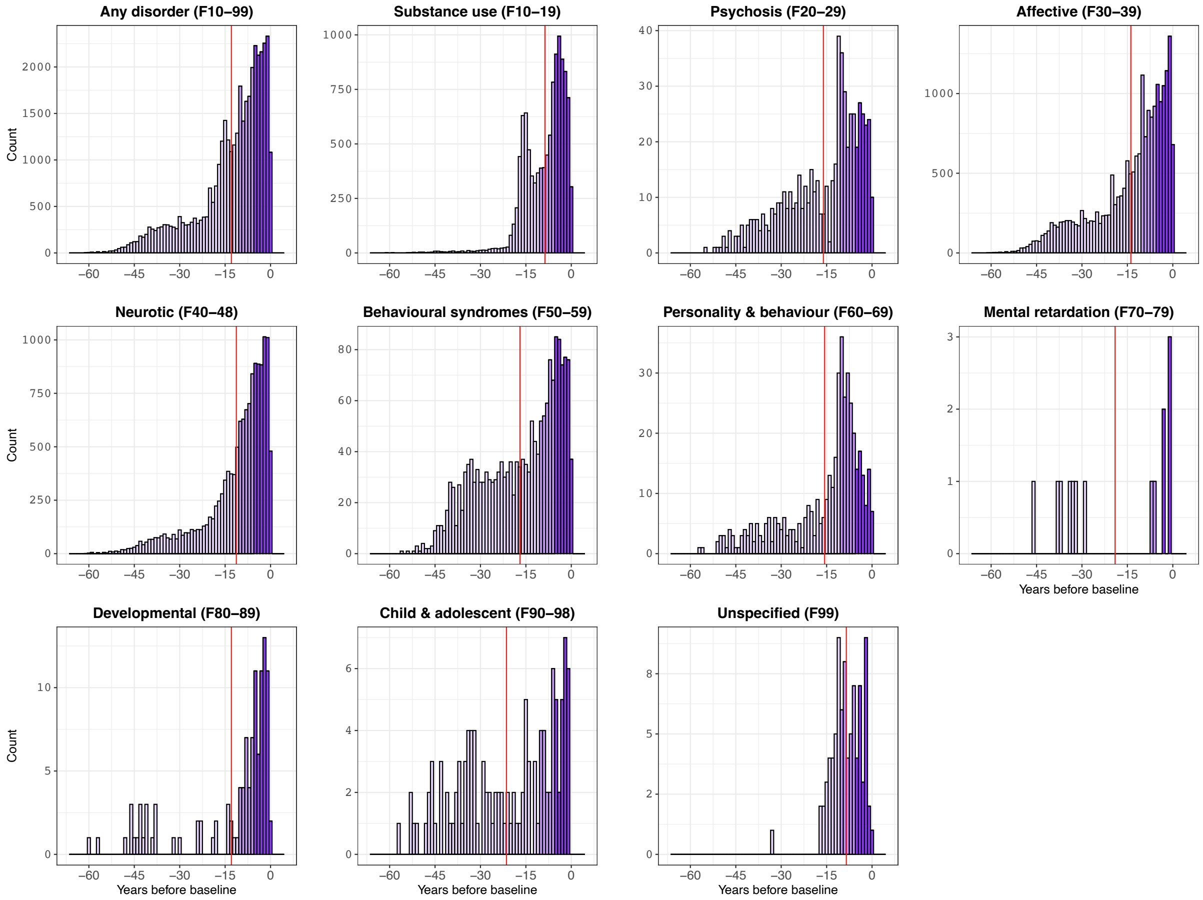

**Figure S3.** Differences in age of onset from the baseline assessment date for groups of two-digit ICD-10 codes for mental and behavioural disorders. ICD-10 = International Classification of Diseases, 10th Revision. Vertical red lines show the mean differences in age of onset. Sample sizes reported in Table 2.

**Table S2.** Age of onset differences from baseline, groups of ICD-10 codes

| ICD-10 | Mean | SD | Median | IQR |
|---|---|---|---|---|
| F10-99, Any disorder | -12.94 | 11.41 | -9.71 | 12.97 |
| F10-19, Substance use | -8.71 | 6.78 | -6.77 | 10.28 |
| F20-29, Psychosis | -16.07 | 12.48 | -11.29 | 18.40 |
| F30-39, Affective | -13.85 | 12.18 | -9.96 | 15.67 |
| F40-48, Neurotic | -11.30 | 10.80 | -7.86 | 11.45 |
| F50-59, Behavioural syndromes | -16.95 | 12.87 | -13.59 | 21.79 |
| F60-69, Personality & behaviour | -15.67 | 12.80 | -10.80 | 14.16 |
| F70-79, Mental retardation | -19.06 | 17.39 | -17.71 | 31.13 |
| F80-89, Developmental | -12.94 | 15.32 | -6.18 | 11.55 |
| F90-98, Child & adolescent | -21.41 | 15.71 | -18.95 | 27.90 |
| F99, Unspecified | -8.50 | 5.12 | -8.57 | 6.84 |

*Note:* ICD-10 = International Classification of Diseases; SD = standard deviation; IQR = interquartile range. Sample sizes reported in Table 2.

### 6. Age of onset differences for individual two-digit ICD-10 codes

**Table S3.** Age of onset differences from baseline, individual ICD-10 codes

| ICD-10 | Mean | SD | Median | IQR |
|---|---|---|---|---|
| F10, Alcohol use | -7.99 | 8.43 | -5.63 | 7.51 |
| F11, Opioid use | -9.22 | 9.85 | -5.93 | 10.05 |
| F12, Cannabis use | -6.49 | 6.90 | -4.33 | 6.48 |
| F13, Sedative use | -7.94 | 7.62 | -4.90 | 8.34 |
| F14, Cocaine use | -6.61 | 4.88 | -6.54 | 3.32 |
| F15, Stimulant use | -7.78 | 8.33 | -5.52 | 6.06 |
| F16, Hallucinogen use | -8.74 | 11.65 | -3.23 | 7.85 |
| F17, Tobacco use | -8.60 | 6.31 | -6.76 | 10.37 |
| F19, Multi-substance use | -16.76 | 12.26 | -14.73 | 18.80 |
| F20, Schizophrenia | -18.33 | 12.49 | -16.28 | 18.75 |
| F22, Delusional | -8.35 | 6.72 | -6.56 | 6.46 |
| F23, Acute psychosis | -10.46 | 11.81 | -7.14 | 8.05 |
| F25, Schizoaffective | -12.06 | 11.71 | -8.61 | 6.66 |
| F29, Psychosis (unspecific) | -11.57 | 10.46 | -8.34 | 13.43 |
| F30, Mania | -14.43 | 11.32 | -10.05 | 16.91 |
| F31, Bipolar | -17.43 | 12.52 | -15.06 | 19.42 |
| F32, Depressive episode | -13.75 | 12.18 | -9.92 | 15.64 |
| F33, Recurrent depression | -8.90 | 7.91 | -7.17 | 6.64 |
| F34, Persistent mood | -11.89 | 11.27 | -8.36 | 13.24 |
| F38, Mood (other) | -6.79 | 4.83 | -5.74 | 4.66 |
| F39, Mood (unspecific) | -6.11 | 6.47 | -4.36 | 5.54 |
| F40, Phobic | -10.38 | 10.43 | -6.59 | 10.73 |
| F41, Anxiety | -12.31 | 11.69 | -8.33 | 13.34 |
| F42, Obsessive-compulsive | -14.84 | 12.31 | -11.01 | 16.95 |
| F43, Stress-related | -7.87 | 6.95 | -6.02 | 8.21 |
| F44, Dissociative | -13.43 | 12.85 | -8.32 | 12.56 |
| F45, Somatoform | -8.78 | 8.29 | -6.33 | 8.68 |
| F48, Other neurotic | -17.92 | 13.46 | -14.74 | 21.35 |
| F50, Eating | -25.96 | 12.50 | -27.52 | 16.54 |
| F51, Sleep | -9.73 | 7.07 | -8.45 | 10.92 |
| F52, Sexual dysfunction | -7.23 | 6.44 | -5.63 | 6.77 |
| F53, Puerperal psychosis | -23.03 | 11.42 | -22.95 | 18.60 |
| F60, Personality | -14.32 | 11.81 | -10.50 | 11.18 |
| F63, Impulse | -19.65 | 16.77 | -15.02 | 26.21 |
| F66, Sexual development | -11.75 | 6.92 | -9.66 | 7.09 |
| F69, Personality (unspecific) | -28.08 | 16.13 | -31.18 | 27.87 |
| F80, Speech and language | -12.50 | 16.60 | -5.79 | 9.20 |
| F81, Scholastic skills | -14.57 | 15.87 | -6.68 | 20.44 |
| F84, Pervasive developmental | -6.21 | 5.07 | -5.61 | 5.37 |
| F90, ADHD | -17.41 | 18.03 | -7.82 | 21.67 |
| F91, Conduct | -18.78 | 14.37 | -15.19 | 24.66 |
| F93, Childhood emotional | -24.15 | 15.35 | -32.58 | 27.45 |
| F95, Tic | -14.74 | 16.54 | -7.52 | 14.34 |
| F98, Child or adolescent (other) | -26.25 | 16.46 | -27.08 | 28.62 |
| F99, Unspecified | -8.50 | 5.12 | -8.57 | 6.84 |

*Note:* ICD-10 = International Classification of Diseases; SD = standard deviation; IQR = interquartile range; ADHD = attention-deficit/hyperactivity disorder. Sample sizes reported in Table S4.

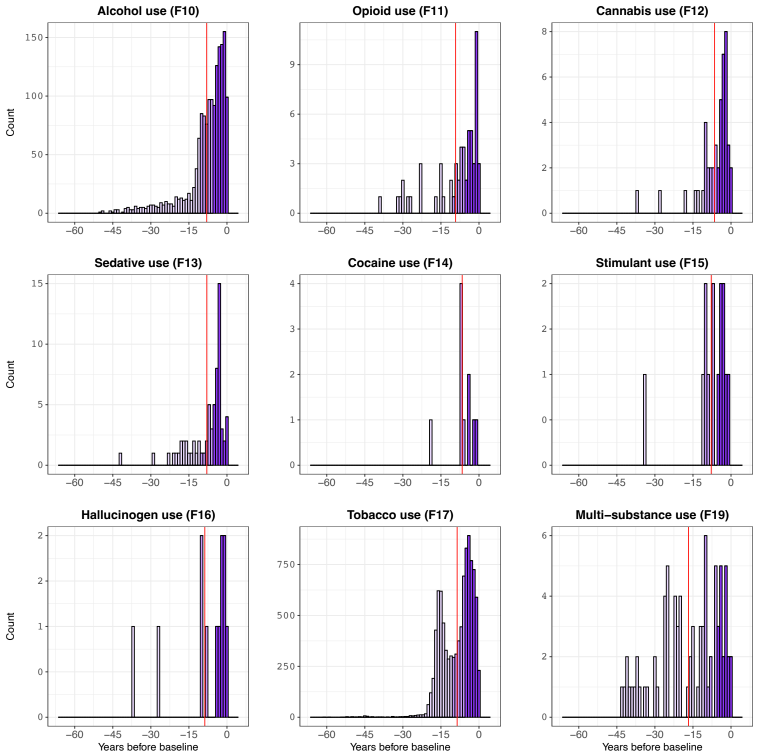

**Figure S4.** Differences in age of onset from the baseline assessment date for individual two-digit ICD-10 codes for mental and behavioural disorders due to psychoactive substance use (F10–F19). ICD-10 = International Classification of Diseases, 10th Revision. Vertical red lines show the mean differences in age of onset. Sample sizes reported in Table S4.

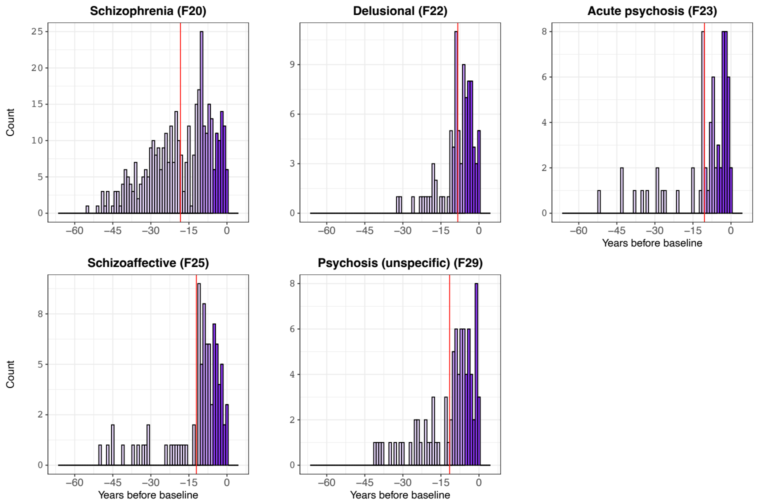

**Figure S5.** Differences in age of onset from the baseline assessment date for individual two-digit ICD-10 codes for schizophrenia, schizotypal and delusional disorders (F20–F29). ICD-10 = International Classification of Diseases, 10th Revision. Vertical red lines show the mean differences in age of onset. Sample sizes reported in Table S4.

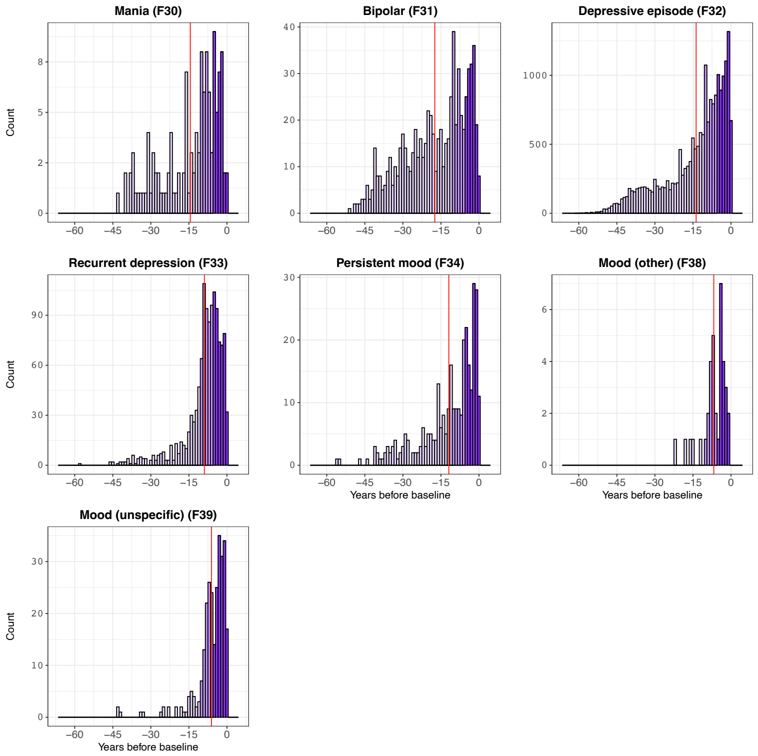

**Figure S6.** Differences in age of onset from the baseline assessment date for individual two-digit ICD-10 codes for mood [affective] disorders (F30–F39). ICD-10 = International Classification of Diseases, 10th Revision. Vertical red lines show the mean differences in age of onset. Sample sizes reported in Table S4.

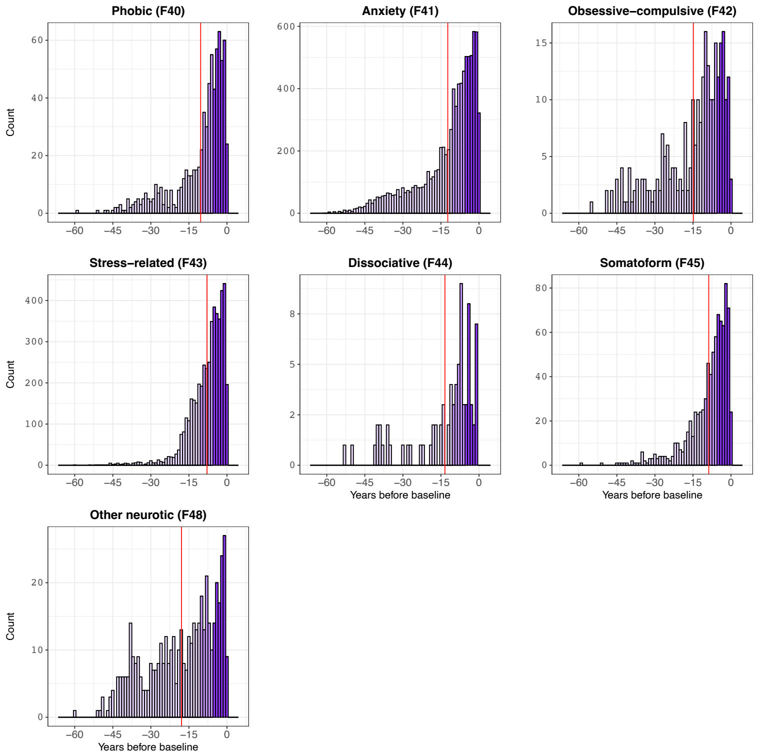

**Figure S7.** Differences in age of onset from the baseline assessment date for individual two-digit ICD-10 codes for neurotic, stress-related and somatoform disorders (F40–F48). ICD-10 = International Classification of Diseases, 10th Revision. Vertical red lines show the mean differences in age of onset. Sample sizes reported in Table S4.

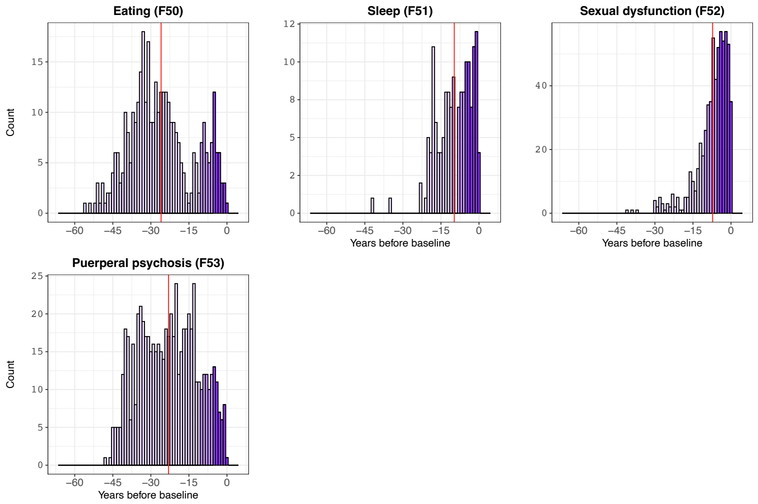

**Figure S8.** Differences in age of onset from the baseline assessment date for individual two-digit ICD-10 codes for behavioural syndromes associated with physiological disturbances and physical factors (F50–F59). ICD-10 = International Classification of Diseases, 10th Revision. Vertical red lines show the mean differences in age of onset. Sample sizes reported in Table S4.

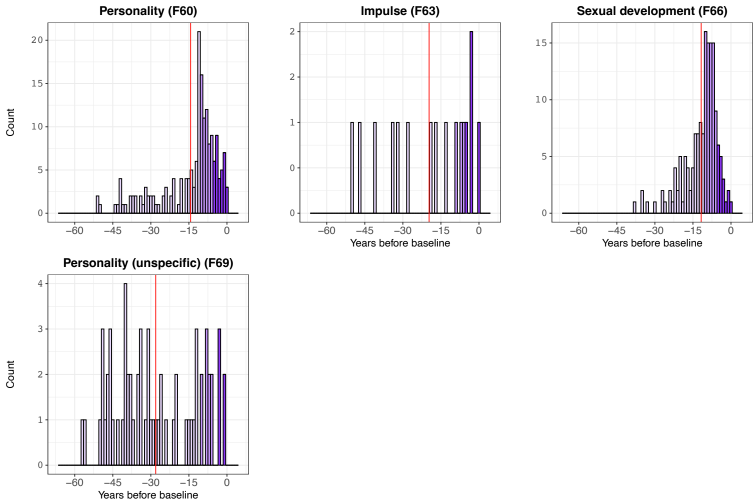

**Figure S9.** Differences in age of onset from the baseline assessment date for individual two-digit ICD-10 codes for disorders of adult personality and behaviour (F60–F69). ICD-10 = International Classification of Diseases, 10th Revision. Vertical red lines show the mean differences in age of onset. Sample sizes reported in Table S4.

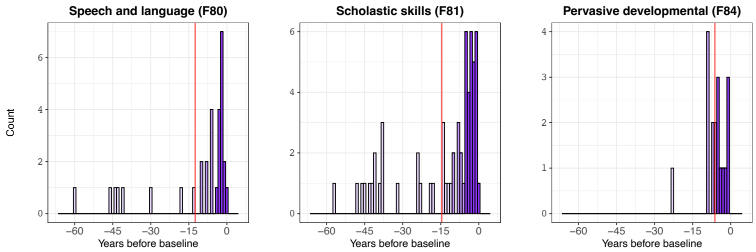

**Figure S10.** Differences in age of onset from the baseline assessment date for individual two-digit ICD-10 codes for disorders of psychological development (F80–F89). ICD-10 = International Classification of Diseases, 10th Revision. Vertical red lines show the mean differences in age of onset. Sample sizes reported in Table S4.

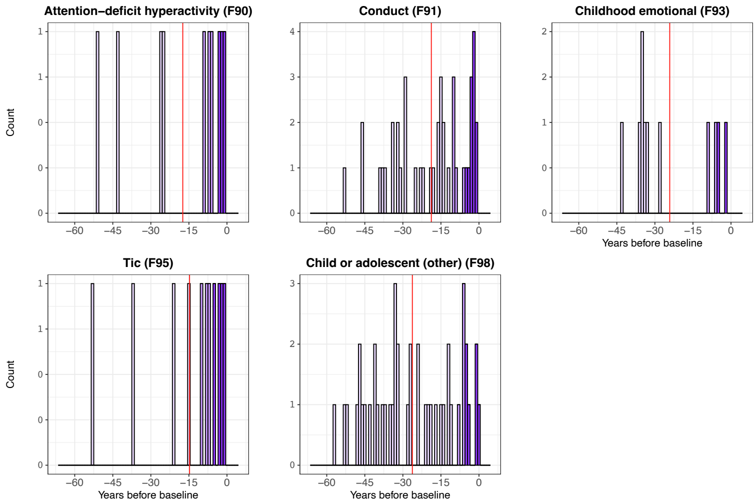

**Figure S11.** Differences in age of onset from the baseline assessment date for individual two-digit ICD-10 codes for disorders of behavioural and emotional disorders with onset usually occurring in childhood and adolescence (F90–F98). ICD-10 = International Classification of Diseases, 10th Revision. Vertical red lines show the mean differences in age of onset. Sample sizes reported in Table S4.

#### 7. MileAge delta and individual two-digit ICD-10 codes

**Table S4.** MileAge delta and individual mental/behavioural disorders

| | | Model 1 (adj. age and sex) | | | | Model 2 (full adjustment) | | | |
|---|---|---|---|---|---|---|---|---|---|
| ICD-10 | *N* | *β* | 95% CI | | *p* | *β* | 95% CI | | *p* |
| None | 186688 | Reference | | | | Reference | | | |
| F10, Alcohol use | 1541 | 0.606 | 0.418 | 0.795 | <0.001 | 0.501 | 0.311 | 0.690 | <0.001 |
| F11, Opioid use | 60 | 1.268 | 0.318 | 2.219 | 0.045 | 1.054 | 0.103 | 2.004 | 0.091 |
| F12, Cannabis use | 46 | -0.914 | -1.999 | 0.172 | 0.255 | -1.108 | -2.193 | -0.023 | 0.127 |
| F13, Sedative use | 68 | -0.394 | -1.287 | 0.498 | 0.580 | -0.479 | -1.371 | 0.413 | 0.501 |
| F14, Cocaine use | 10 | 0.965 | -1.363 | 3.293 | 0.604 | 0.710 | -1.616 | 3.036 | 0.722 |
| F15, Stimulant use | 14 | 0.942 | -1.026 | 2.909 | 0.570 | 0.739 | -1.227 | 2.705 | 0.633 |
| F16, Hallucinogen use | 12 | 3.758 | 1.633 | 5.883 | 0.005 | 3.630 | 1.507 | 5.753 | 0.007 |
| F17, Tobacco use | 9715 | 0.409 | 0.332 | 0.485 | <0.001 | 0.373 | 0.296 | 0.450 | <0.001 |
| F19, Multi-substance use | 81 | 0.624 | -0.194 | 1.442 | 0.338 | 0.461 | -0.357 | 1.278 | 0.493 |
| F20, Schizophrenia | 386 | 0.859 | 0.484 | 1.235 | <0.001 | 0.638 | 0.261 | 1.014 | 0.007 |
| F22, Delusional | 88 | 0.956 | 0.171 | 1.741 | 0.065 | 0.804 | 0.020 | 1.589 | 0.127 |
| F23, Acute psychosis | 66 | 0.044 | -0.862 | 0.950 | 0.956 | -0.069 | -0.974 | 0.837 | 0.937 |
| F25, Schizoaffective | 85 | 0.905 | 0.106 | 1.703 | 0.085 | 0.729 | -0.069 | 1.527 | 0.200 |
| F29, Psychosis (unspecific) | 83 | 0.541 | -0.267 | 1.349 | 0.426 | 0.366 | -0.442 | 1.174 | 0.574 |
| F30, Mania | 124 | 0.494 | -0.167 | 1.155 | 0.348 | 0.408 | -0.252 | 1.069 | 0.474 |
| F31, Bipolar | 711 | 0.233 | -0.044 | 0.509 | 0.255 | 0.146 | -0.131 | 0.423 | 0.501 |
| F32, Depressive episode | 19176 | 0.274 | 0.218 | 0.331 | <0.001 | 0.230 | 0.173 | 0.286 | <0.001 |
| F33, Recurrent depression | 1211 | 0.327 | 0.115 | 0.539 | 0.015 | 0.268 | 0.056 | 0.480 | 0.055 |
| F34, Persistent mood | 306 | 0.247 | -0.174 | 0.668 | 0.480 | 0.190 | -0.231 | 0.611 | 0.574 |
| F38, Mood (other) | 36 | 0.300 | -0.927 | 1.527 | 0.758 | 0.237 | -0.989 | 1.462 | 0.824 |
| F39, Mood (unspecific) | 282 | 0.201 | -0.238 | 0.640 | 0.574 | 0.171 | -0.268 | 0.609 | 0.633 |
| F40, Phobic | 717 | 0.151 | -0.125 | 0.426 | 0.500 | 0.119 | -0.156 | 0.395 | 0.584 |
| F41, Anxiety | 8433 | 0.131 | 0.049 | 0.213 | 0.013 | 0.098 | 0.016 | 0.180 | 0.070 |
| F42, Obsessive-compulsive | 270 | -0.518 | -0.966 | -0.070 | 0.079 | -0.588 | -1.036 | -0.140 | 0.045 |
| F43, Stress-related | 4749 | 0.193 | 0.084 | 0.301 | 0.005 | 0.169 | 0.061 | 0.277 | 0.014 |
| F44, Dissociative | 79 | 0.374 | -0.454 | 1.203 | 0.574 | 0.317 | -0.510 | 1.145 | 0.633 |
| F45, Somatoform | 843 | 0.021 | -0.233 | 0.276 | 0.937 | 0.017 | -0.237 | 0.271 | 0.937 |
| F48, Other neurotic | 487 | 0.101 | -0.233 | 0.435 | 0.722 | 0.038 | -0.296 | 0.372 | 0.925 |
| F50, Eating | 364 | -0.588 | -0.975 | -0.201 | 0.016 | -0.613 | -1.000 | -0.227 | 0.013 |
| F51, Sleep | 153 | 0.367 | -0.228 | 0.963 | 0.474 | 0.362 | -0.232 | 0.957 | 0.475 |
| F52, Sexual dysfunction | 632 | -0.008 | -0.301 | 0.286 | 0.971 | -0.012 | -0.305 | 0.281 | 0.959 |
| F53, Puerperal psychosis | 629 | -0.389 | -0.683 | -0.094 | 0.045 | -0.371 | -0.666 | -0.077 | 0.055 |
| F60, Personality | 180 | 0.805 | 0.256 | 1.354 | 0.021 | 0.667 | 0.118 | 1.216 | 0.065 |
| F63, Impulse | 16 | 1.142 | -0.698 | 2.982 | 0.474 | 1.083 | -0.755 | 2.922 | 0.480 |
| F66, Sexual development | 155 | -0.333 | -0.924 | 0.259 | 0.493 | -0.351 | -0.942 | 0.240 | 0.480 |
| F69, Personality (unspecific) | 64 | -0.003 | -0.923 | 0.918 | 0.996 | -0.089 | -1.009 | 0.830 | 0.932 |
| F80, Speech and language | 31 | 0.441 | -0.881 | 1.763 | 0.690 | 0.375 | -0.946 | 1.696 | 0.722 |
| F81, Scholastic skills | 60 | 0.355 | -0.596 | 1.305 | 0.633 | 0.236 | -0.714 | 1.186 | 0.758 |
| F84, Pervasive developmental | 18 | 0.497 | -1.238 | 2.232 | 0.722 | 0.357 | -1.376 | 2.091 | 0.813 |
| F90, ADHD | 10 | -0.324 | -2.652 | 2.003 | 0.894 | -0.373 | -2.699 | 1.952 | 0.869 |
| F91, Conduct | 43 | 1.314 | 0.192 | 2.437 | 0.075 | 1.238 | 0.117 | 2.360 | 0.091 |
| F93, Childhood emotional | 11 | 0.650 | -1.570 | 2.869 | 0.722 | 0.594 | -1.624 | 2.811 | 0.739 |
| F95, Tic | 11 | 1.550 | -0.669 | 3.770 | 0.397 | 1.544 | -0.673 | 3.761 | 0.397 |
| F98, Child or adolescent (other) | 45 | -0.580 | -1.678 | 0.517 | 0.501 | -0.612 | -1.709 | 0.484 | 0.493 |
| F99, Unspecified | 86 | 0.085 | -0.708 | 0.879 | 0.925 | -0.059 | -0.852 | 0.735 | 0.937 |

*Note:* ICD-10 = International Classification of Diseases, 10th Revision; CI = confidence interval. Model 1–adjusted for chronological age and sex; Model 2–adjusted for chronological age, sex, ethnicity, cohabitation with spouse/partner, highest educational/professional qualification, annual gross household income, Townsend deprivation index and fasting time. Cells highlighted in grey and blue correspond to statistically significant associations (nominally and after multiple testing corrections, respectively). *P*-values shown are corrected for multiple testing using the Benjamini–Hochberg procedure.

### 8. MileAge delta and groups of two-digit ICD-10 codes; sex-stratified

**Table S5.** MileAge delta and groups of mental/behavioural disorders stratified by sex

| | Males | | | | | | | | | Females | | | | | | | | |
|---|---|---|---|---|---|---|---|---|---|---|---|---|---|---|---|---|---|---|
| | | Model 1 (adj. age and sex) | | | | Model 2 (full adjustment) | | | | | Model 1 | | | | Model 2 | | | |
| ICD-10 | *N* | *β* | 95% CI | | *p* | *β* | 95% CI | | *p* | *N* | *β* | 95% CI | | *p* | *β* | 95% CI | | *p* |
| None | 88232 | Reference | | | | Reference | | | | 98456 | Reference | | | | Reference | | | |
| F10-99, Any disorder | 15449 | 0.362 | 0.298 | 0.426 | <0.001 | 0.313 | 0.248 | 0.377 | <0.001 | 23075 | 0.188 | 0.134 | 0.242 | <0.001 | 0.163 | 0.109 | 0.218 | <0.001 |
| F10-19, Substance use | 6040 | 0.524 | 0.427 | 0.621 | <0.001 | 0.467 | 0.369 | 0.566 | <0.001 | 5121 | 0.350 | 0.244 | 0.456 | <0.001 | 0.324 | 0.218 | 0.430 | <0.001 |
| F20-29, Psychosis | 343 | 0.967 | 0.572 | 1.363 | <0.001 | 0.765 | 0.368 | 1.163 | <0.001 | 244 | 0.350 | -0.123 | 0.823 | 0.215 | 0.239 | -0.235 | 0.712 | 0.431 |
| F30-39, Affective | 6992 | 0.358 | 0.267 | 0.449 | <0.001 | 0.300 | 0.208 | 0.392 | <0.001 | 13368 | 0.238 | 0.170 | 0.306 | <0.001 | 0.204 | 0.135 | 0.272 | <0.001 |
| F40-48, Neurotic | 4938 | 0.209 | 0.102 | 0.316 | <0.001 | 0.175 | 0.068 | 0.282 | 0.003 | 8978 | 0.136 | 0.055 | 0.217 | 0.003 | 0.115 | 0.033 | 0.196 | 0.013 |
| F50-59, Behavioural syndromes | 459 | 0.413 | 0.071 | 0.755 | 0.036 | 0.393 | 0.052 | 0.735 | 0.044 | 1304 | -0.309 | -0.515 | -0.103 | 0.008 | -0.312 | -0.518 | -0.106 | 0.008 |
| F60-69, Personality & behaviour | 250 | 0.121 | -0.342 | 0.584 | 0.788 | 0.042 | -0.421 | 0.504 | 0.911 | 183 | 0.608 | 0.062 | 1.154 | 0.058 | 0.550 | 0.004 | 1.096 | 0.089 |
| F70-79, Mental retardation | | *N* insufficient | | | | | | | | | *N* insufficient | | | | | | | |
| F80-89, Developmental | 68 | 0.408 | -0.479 | 1.294 | 0.539 | 0.290 | -0.597 | 1.176 | 0.717 | 47 | 0.405 | -0.671 | 1.482 | 0.533 | 0.317 | -0.759 | 1.393 | 0.620 |
| F90-98, Child & adolescent | 66 | 0.135 | -0.764 | 1.035 | 0.894 | 0.075 | -0.824 | 0.974 | 0.911 | 54 | 0.883 | -0.121 | 1.887 | 0.144 | 0.846 | -0.157 | 1.849 | 0.155 |
| F99, Unspecified | 35 | 0.183 | -1.053 | 1.418 | 0.894 | 0.001 | -1.233 | 1.236 | 0.998 | 51 | 0.068 | -0.965 | 1.101 | 0.940 | -0.032 | -1.065 | 1.001 | 0.952 |

*Note:* ICD-10 = International Classification of Diseases, 10th Revision; CI = confidence interval. Model 1–adjusted for chronological age; Model 2–adjusted for chronological age, ethnicity, cohabitation with spouse/partner, highest educational/professional qualification, annual gross household income, Townsend deprivation index and fasting time. Cells highlighted in grey and blue correspond to statistically significant associations (nominally and after multiple testing corrections, respectively). *P*-values shown are corrected for multiple testing using the Benjamini–Hochberg procedure within each sex.

### 9. MileAge delta and individual two-digit ICD-10 codes; sex-stratified

**Table S6.** MileAge delta and individual mental/behavioural disorders stratified by sex

| | Males | | | | | | | | | Females | | | | | | | |
|---|---|---|---|---|---|---|---|---|---|---|---|---|---|---|---|---|---|
| | | Model 1 (adj. age and sex) | | | | Model 2 (full adjustment) | | | | | Model 1 | | | | Model 2 | | |
| ICD-10 | *N* | *β* | 95% CI | | *p* | *β* | 95% CI | | *p* | *N* | *β* | 95% CI | | *p* | *β* | 95% CI | | *p* |
| None | 88232 | Reference | | | | Reference | | | | 98456 | Reference | | | | Reference | | | |
| F10, Alcohol use | 1134 | 0.468 | 0.249 | 0.687 | <0.001 | 0.367 | 0.146 | 0.587 | 0.009 | 407 | 1.008 | 0.641 | 1.374 | <0.001 | 0.938 | 0.571 | 1.306 | <0.001 |
| F11, Opioid use | 44 | 1.376 | 0.275 | 2.478 | 0.076 | 1.139 | 0.037 | 2.242 | 0.151 | 16 | 0.613 | -1.231 | 2.457 | 0.786 | 0.562 | -1.281 | 2.405 | 0.786 |
| F12, Cannabis use | 34 | -1.523 | -2.776 | -0.270 | 0.085 | -1.688 | -2.941 | -0.434 | 0.056 | 12 | 0.521 | -1.609 | 2.651 | 0.818 | 0.323 | -1.806 | 2.451 | 0.841 |
| F13, Sedative use | 24 | -1.511 | -3.003 | -0.020 | 0.151 | -1.686 | -3.177 | -0.196 | 0.109 | 44 | 0.186 | -0.926 | 1.298 | 0.841 | 0.158 | -0.953 | 1.269 | 0.841 |
| F14, Cocaine use | | *N* insufficient | | | | | | | | | *N* insufficient | | | | | | | |
| F15, Stimulant use | 10 | 1.294 | -1.017 | 3.604 | 0.403 | 1.081 | -1.228 | 3.390 | 0.492 | | *N* insufficient | | | | | | | |
| F16, Hallucinogen use | | *N* insufficient | | | | | | | | | *N* insufficient | | | | | | | |
| F17, Tobacco use | 4997 | 0.535 | 0.429 | 0.641 | <0.001 | 0.488 | 0.381 | 0.595 | <0.001 | 4718 | 0.308 | 0.198 | 0.418 | <0.001 | 0.286 | 0.176 | 0.397 | <0.001 |
| F19, Multi-substance use | 50 | 0.367 | -0.666 | 1.401 | 0.643 | 0.186 | -0.847 | 1.220 | 0.864 | 31 | 0.869 | -0.456 | 2.194 | 0.440 | 0.780 | -0.545 | 2.104 | 0.497 |
| F20, Schizophrenia | 253 | 1.043 | 0.583 | 1.503 | <0.001 | 0.807 | 0.345 | 1.269 | 0.006 | 133 | 0.343 | -0.297 | 0.983 | 0.573 | 0.204 | -0.436 | 0.845 | 0.786 |
| F22, Delusional | 41 | 1.521 | 0.379 | 2.662 | 0.056 | 1.363 | 0.222 | 2.504 | 0.087 | 47 | 0.507 | -0.569 | 1.583 | 0.656 | 0.391 | -0.685 | 1.466 | 0.786 |
| F23, Acute psychosis | 29 | 1.209 | -0.148 | 2.566 | 0.239 | 1.105 | -0.252 | 2.461 | 0.296 | 37 | -0.797 | -2.010 | 0.416 | 0.440 | -0.889 | -2.101 | 0.324 | 0.395 |
| F25, Schizoaffective | 43 | 1.322 | 0.208 | 2.437 | 0.087 | 1.131 | 0.017 | 2.245 | 0.151 | 42 | 0.453 | -0.686 | 1.591 | 0.753 | 0.339 | -0.799 | 1.477 | 0.786 |
| F29, Psychosis (unspecific) | 40 | 0.590 | -0.565 | 1.746 | 0.442 | 0.401 | -0.754 | 1.556 | 0.644 | 43 | 0.467 | -0.658 | 1.592 | 0.735 | 0.356 | -0.768 | 1.481 | 0.786 |
| F30, Mania | 52 | -0.048 | -1.061 | 0.966 | 0.939 | -0.163 | -1.176 | 0.850 | 0.865 | 72 | 0.897 | 0.027 | 1.766 | 0.168 | 0.830 | -0.039 | 1.699 | 0.202 |
| F31, Bipolar | 293 | -0.137 | -0.565 | 0.291 | 0.677 | -0.228 | -0.656 | 0.200 | 0.423 | 418 | 0.537 | 0.175 | 0.898 | 0.040 | 0.467 | 0.105 | 0.829 | 0.062 |
| F32, Depressive episode | 6574 | 0.390 | 0.296 | 0.483 | <0.001 | 0.332 | 0.238 | 0.426 | <0.001 | 12602 | 0.232 | 0.162 | 0.302 | <0.001 | 0.197 | 0.127 | 0.268 | <0.001 |
| F33, Recurrent depression | 412 | 0.355 | -0.006 | 0.715 | 0.167 | 0.290 | -0.071 | 0.651 | 0.296 | 799 | 0.341 | 0.078 | 0.603 | 0.062 | 0.297 | 0.034 | 0.559 | 0.119 |
| F34, Persistent mood | 102 | 0.511 | -0.213 | 1.235 | 0.342 | 0.459 | -0.265 | 1.182 | 0.368 | 204 | 0.183 | -0.334 | 0.700 | 0.786 | 0.121 | -0.396 | 0.638 | 0.818 |
| F38, Mood (other) | | *N* insufficient | | | | | | | | 27 | 0.709 | -0.711 | 2.129 | 0.623 | 0.659 | -0.760 | 2.078 | 0.656 |
| F39, Mood (unspecific) | 60 | 0.762 | -0.182 | 1.705 | 0.296 | 0.694 | -0.248 | 1.637 | 0.342 | 222 | 0.144 | -0.352 | 0.640 | 0.786 | 0.124 | -0.372 | 0.619 | 0.818 |
| F40, Phobic | 164 | 0.458 | -0.113 | 1.029 | 0.296 | 0.405 | -0.165 | 0.976 | 0.342 | 553 | 0.052 | -0.263 | 0.366 | 0.841 | 0.028 | -0.287 | 0.342 | 0.886 |
| F41, Anxiety | 3009 | 0.139 | 0.003 | 0.274 | 0.151 | 0.096 | -0.040 | 0.232 | 0.342 | 5424 | 0.142 | 0.039 | 0.245 | 0.052 | 0.121 | 0.018 | 0.224 | 0.109 |
| F42, Obsessive-compulsive | 120 | -0.186 | -0.853 | 0.482 | 0.735 | -0.264 | -0.931 | 0.404 | 0.590 | 150 | -0.796 | -1.398 | -0.193 | 0.061 | -0.847 | -1.450 | -0.245 | 0.049 |
| F43, Stress-related | 1670 | 0.313 | 0.132 | 0.493 | 0.006 | 0.290 | 0.110 | 0.471 | 0.012 | 3079 | 0.155 | 0.020 | 0.290 | 0.117 | 0.130 | -0.005 | 0.266 | 0.202 |
| F44, Dissociative | 28 | -0.119 | -1.500 | 1.262 | 0.890 | -0.154 | -1.534 | 1.226 | 0.887 | 51 | 0.731 | -0.302 | 1.764 | 0.419 | 0.669 | -0.364 | 1.701 | 0.440 |
| F45, Somatoform | 293 | -0.037 | -0.465 | 0.390 | 0.890 | -0.049 | -0.476 | 0.378 | 0.887 | 550 | 0.046 | -0.269 | 0.361 | 0.841 | 0.047 | -0.268 | 0.362 | 0.841 |
| F48, Other neurotic | 201 | 0.372 | -0.144 | 0.888 | 0.342 | 0.302 | -0.214 | 0.817 | 0.396 | 286 | -0.061 | -0.498 | 0.375 | 0.841 | -0.107 | -0.543 | 0.330 | 0.818 |
| F50, Eating | 22 | 1.079 | -0.479 | 2.637 | 0.349 | 1.016 | -0.540 | 2.573 | 0.368 | 342 | -0.538 | -0.938 | -0.138 | 0.058 | -0.572 | -0.972 | -0.173 | 0.048 |
| F51, Sleep | 63 | 0.597 | -0.324 | 1.517 | 0.368 | 0.604 | -0.316 | 1.523 | 0.368 | 90 | 0.235 | -0.543 | 1.013 | 0.786 | 0.227 | -0.551 | 1.004 | 0.786 |
| F52, Sexual dysfunction | 368 | 0.301 | -0.081 | 0.683 | 0.302 | 0.281 | -0.100 | 0.662 | 0.342 | 264 | -0.292 | -0.746 | 0.163 | 0.440 | -0.286 | -0.740 | 0.168 | 0.447 |
| F53, Puerperal psychosis | | *N* insufficient | | | | | | | | 627 | -0.287 | -0.583 | 0.009 | 0.202 | -0.276 | -0.572 | 0.020 | 0.213 |
| F60, Personality | 78 | 1.070 | 0.242 | 1.898 | 0.064 | 0.906 | 0.078 | 1.734 | 0.124 | 102 | 0.657 | -0.073 | 1.388 | 0.237 | 0.576 | -0.155 | 1.307 | 0.333 |
| F63, Impulse | | *N* insufficient | | | | | | | | 10 | 2.075 | -0.258 | 4.407 | 0.238 | 2.017 | -0.315 | 4.348 | 0.253 |
| F66, Sexual development | 111 | -0.396 | -1.090 | 0.299 | 0.403 | -0.430 | -1.123 | 0.264 | 0.375 | 44 | 0.152 | -0.960 | 1.264 | 0.841 | 0.146 | -0.965 | 1.257 | 0.841 |

| | | | | | | | | | | | | | | | | | | |
|---|---|---|---|---|---|---|---|---|---|---|---|---|---|---|---|---|---|---|
| F69, Personality (unspecific) | 33 | -0.215 | -1.487 | 1.057 | 0.865 | -0.273 | -1.544 | 0.998 | 0.817 | 31 | 0.261 | -1.064 | 1.586 | 0.841 | 0.157 | -1.168 | 1.481 | 0.850 |
| F80, Speech and language | 21 | -0.141 | -1.736 | 1.453 | 0.890 | -0.185 | -1.778 | 1.408 | 0.887 | 10 | 1.598 | -0.734 | 3.931 | 0.440 | 1.516 | -0.815 | 3.847 | 0.440 |
| F81, Scholastic skills | 29 | 0.338 | -1.019 | 1.695 | 0.772 | 0.209 | -1.148 | 1.566 | 0.865 | 31 | 0.408 | -0.917 | 1.733 | 0.786 | 0.319 | -1.006 | 1.645 | 0.818 |
| F84, Pervasive developmental | 15 | 1.203 | -0.684 | 3.089 | 0.368 | 1.003 | -0.883 | 2.888 | 0.423 | | *N* insufficient | | | | | | | |
| F90, ADHD | | *N* insufficient | | | | | | | | | *N* insufficient | | | | | | | |
| F91, Conduct | 29 | 0.824 | -0.533 | 2.181 | 0.376 | 0.765 | -0.591 | 2.121 | 0.403 | 14 | 2.070 | 0.099 | 4.042 | 0.167 | 2.022 | 0.052 | 3.992 | 0.168 |
| F93, Childhood emotional | | *N* insufficient | | | | | | | | | *N* insufficient | | | | | | | |
| F95, Tic | | *N* insufficient | | | | | | | | | *N* insufficient | | | | | | | |
| F98, Child or adolescent (other) | 26 | -0.882 | -2.315 | 0.551 | 0.375 | -0.940 | -2.371 | 0.492 | 0.368 | 19 | -0.298 | -1.991 | 1.394 | 0.841 | -0.322 | -2.013 | 1.369 | 0.841 |
| F99, Unspecified | 35 | 0.183 | -1.053 | 1.418 | 0.865 | 0.001 | -1.233 | 1.236 | 0.998 | 51 | 0.068 | -0.965 | 1.101 | 0.909 | -0.032 | -1.065 | 1.001 | 0.952 |

*Note:* ICD-10 = International Classification of Diseases, 10th Revision; ADHD = attention-deficit/hyperactivity disorder; CI = confidence interval. Model 1–adjusted for chronological age; Model 2–adjusted for chronological age, ethnicity, cohabitation with spouse/partner, highest educational/professional qualification, annual gross household income, Townsend deprivation index and fasting time. Cells highlighted in grey and blue correspond to statistically significant associations (nominally and after multiple testing corrections, respectively). *P*-values shown are corrected for multiple testing using the Benjamini–Hochberg procedure within each sex.

#### 10. MileAge delta and polygenic scores for groups of disorders

**Table S7.** MileAge delta and polygenic scores for groups of disorders across ancestries

| Polygenic score | *β* | 95% CI | | *p* |
|---|---|---|---|---|
| F10-99, Any disorder | 0.003 | -0.013 | 0.020 | 0.878 |
| F10-19, Substance use | 0.018 | 0.002 | 0.034 | 0.141 |
| F20-29, Psychosis | -0.036 | -0.052 | -0.020 | <0.001 |
| F30-39, Affective | 0.008 | -0.009 | 0.024 | 0.762 |
| F40-48, Neurotic | -0.001 | -0.018 | 0.016 | 0.974 |
| F50-59, Behavioural syndromes | 0.000 | -0.017 | 0.016 | 0.974 |
| F70-79, Mental retardation | 0.007 | -0.010 | 0.023 | 0.762 |
| F80-89, Developmental | 0.009 | -0.008 | 0.025 | 0.762 |
| F90-98, Child & adolescent | 0.004 | -0.012 | 0.019 | 0.878 |

*Note:* CI = confidence interval. Models were adjusted for chronological age, sex, assessment centre, batch number, the first six genetic principal components and fasting time. Cells highlighted in grey and blue correspond to statistically significant associations (nominally and after multiple testing corrections, respectively). *P*-values shown are corrected for multiple testing using the Benjamini–Hochberg procedure. $N$ = 219,494.

**Table S8.** Ancestry-specific associations between MileAge delta and polygenic scores for groups of disorders

| Polygenic score | Population | *β* | 95% CI | | *p* |
|---|---|---|---|---|---|
| F10-99, Any disorder | AFR | 0.041 | -0.102 | 0.183 | 0.891 |
| | AMR | 0.601 | 0.065 | 1.136 | 0.127 |
| | CSA | 0.038 | -0.092 | 0.167 | 0.733 |
| | EAS | -0.097 | -0.388 | 0.195 | 0.662 |
| | EUR | 0.002 | -0.016 | 0.019 | 0.967 |
| | MID | 0.205 | -0.838 | 1.248 | 0.783 |
| F10-19, Substance use | AFR | -0.014 | -0.159 | 0.131 | 0.891 |
| | AMR | 1.011 | 0.354 | 1.669 | 0.025 |
| | CSA | 0.071 | -0.058 | 0.201 | 0.631 |
| | EAS | 0.165 | -0.121 | 0.452 | 0.637 |
| | EUR | 0.016 | -0.001 | 0.032 | 0.290 |
| | MID | -0.504 | -1.596 | 0.587 | 0.783 |
| F20-29, Psychosis | AFR | -0.009 | -0.140 | 0.122 | 0.891 |
| | AMR | 0.060 | -0.641 | 0.760 | 0.867 |
| | CSA | -0.097 | -0.232 | 0.038 | 0.553 |
| | EAS | 0.133 | -0.148 | 0.414 | 0.637 |
| | EUR | -0.038 | -0.055 | -0.021 | <0.001 |
| | MID | 0.705 | -0.290 | 1.700 | 0.728 |
| F30-39, Affective | AFR | -0.038 | -0.185 | 0.109 | 0.891 |
| | AMR | 0.448 | -0.175 | 1.071 | 0.237 |
| | CSA | 0.092 | -0.044 | 0.228 | 0.553 |
| | EAS | -0.166 | -0.453 | 0.121 | 0.637 |
| | EUR | 0.007 | -0.010 | 0.024 | 0.794 |
| | MID | 0.115 | -0.986 | 1.215 | 0.836 |
| F40-48, Neurotic | AFR | 0.027 | -0.118 | 0.172 | 0.891 |
| | AMR | 0.533 | -0.098 | 1.165 | 0.175 |
| | CSA | 0.128 | -0.008 | 0.264 | 0.553 |
| | EAS | -0.105 | -0.400 | 0.191 | 0.662 |
| | EUR | -0.003 | -0.020 | 0.014 | 0.891 |
| | MID | 0.319 | -0.690 | 1.328 | 0.783 |
| F50-59, Behavioural syndromes | AFR | 0.022 | -0.109 | 0.154 | 0.891 |
| | AMR | 0.573 | -0.074 | 1.220 | 0.175 |
| | CSA | 0.014 | -0.121 | 0.149 | 0.870 |
| | EAS | -0.336 | -0.656 | -0.016 | 0.357 |
| | EUR | 0.000 | -0.017 | 0.017 | 0.992 |
| | MID | 0.875 | -0.202 | 1.953 | 0.728 |
| F70-79, Mental retardation | AFR | -0.030 | -0.181 | 0.122 | 0.891 |
| | AMR | -0.325 | -1.077 | 0.427 | 0.507 |
| | CSA | 0.052 | -0.088 | 0.192 | 0.733 |
| | EAS | 0.024 | -0.277 | 0.325 | 0.878 |
| | EUR | 0.007 | -0.010 | 0.023 | 0.794 |
| | MID | -0.210 | -1.140 | 0.720 | 0.783 |
| F80-89, Developmental | AFR | 0.023 | -0.126 | 0.172 | 0.891 |
| | AMR | 0.075 | -0.539 | 0.690 | 0.867 |
| | CSA | 0.011 | -0.117 | 0.139 | 0.870 |
| | EAS | 0.042 | -0.249 | 0.333 | 0.874 |
| | EUR | 0.008 | -0.009 | 0.024 | 0.794 |
| | MID | -0.313 | -1.125 | 0.498 | 0.783 |
| F90-98, Child & adolescent | AFR | -0.066 | -0.216 | 0.084 | 0.891 |
| | AMR | 0.628 | -0.041 | 1.296 | 0.175 |
| | CSA | 0.040 | -0.089 | 0.169 | 0.733 |
| | EAS | -0.149 | -0.423 | 0.125 | 0.637 |
| | EUR | 0.004 | -0.012 | 0.020 | 0.891 |
| | MID | 0.308 | -0.771 | 1.388 | 0.783 |

*Note:* AFR = African; AMR = Admixed American; EAS = East Asian; EUR = European; CSA = Central and South Asian; MID = Middle Eastern. Models were adjusted for chronological age, sex, assessment centre, batch number (except for MID), the first six genetic principal components and fasting time. Cells highlighted in grey and blue correspond to statistically significant associations (nominally and after multiple testing corrections, respectively). *P*-values shown are corrected for multiple testing using the Benjamini–Hochberg procedure (separately within each population). *N* = 3426 (AFR); *N* = 278 (AMR); *N* = 3916 (CSA); *N* = 1012 (EAS); *N* = 210,755 (EUR); *N* = 94 (MID).

### 11. MileAge delta and polygenic scores for individual disorders

**Table S9.** MileAge delta and polygenic scores for individual disorders across ancestries

| Polygenic score | *β* | 95% CI | | *p* |
|---|---|---|---|---|
| F10, Alcohol use | -0.010 | -0.027 | 0.007 | 0.448 |
| F11, Opioid use | -0.009 | -0.026 | 0.009 | 0.499 |
| F12, Cannabis use | 0.001 | -0.015 | 0.018 | 0.922 |
| F13, Sedative use | -0.002 | -0.019 | 0.014 | 0.886 |
| F15, Stimulant use | -0.003 | -0.020 | 0.014 | 0.886 |
| F17, Tobacco use | -0.034 | -0.051 | -0.017 | <0.001 |
| F19, Multi-substance use | 0.000 | -0.017 | 0.017 | 0.991 |
| Schizophrenia | -0.009 | -0.024 | 0.007 | 0.448 |
| F22, Delusional | -0.019 | -0.035 | -0.004 | 0.053 |
| F23, Acute psychosis | -0.040 | -0.056 | -0.023 | <0.001 |
| F25, Schizoaffective | -0.023 | -0.038 | -0.007 | 0.022 |
| F29, Psychosis (unspecific) | -0.027 | -0.043 | -0.011 | 0.008 |
| F30, Mania | -0.003 | -0.020 | 0.014 | 0.886 |
| Bipolar | -0.002 | -0.017 | 0.013 | 0.886 |
| F32, Depressive episode | 0.011 | -0.005 | 0.028 | 0.417 |
| F33, Recurrent depression | 0.004 | -0.012 | 0.021 | 0.799 |
| Depression | 0.020 | 0.004 | 0.035 | 0.048 |
| F34, Persistent mood | -0.012 | -0.028 | 0.005 | 0.403 |
| F39, Mood (unspecific) | 0.013 | -0.003 | 0.030 | 0.357 |
| F40, Phobic | -0.011 | -0.029 | 0.006 | 0.435 |
| F41, Anxiety | -0.009 | -0.024 | 0.007 | 0.448 |
| F42, Obsessive-compulsive | -0.035 | -0.052 | -0.019 | <0.001 |
| F43, Stress-related | -0.009 | -0.026 | 0.008 | 0.448 |
| F44, Dissociative | 0.004 | -0.012 | 0.021 | 0.799 |
| F45, Somatoform | -0.012 | -0.028 | 0.005 | 0.403 |
| F48, Other neurotic | -0.009 | -0.025 | 0.006 | 0.448 |
| F50, Eating | 0.001 | -0.016 | 0.017 | 0.964 |
| Anorexia | -0.023 | -0.038 | -0.008 | 0.013 |
| F51, Sleep | -0.007 | -0.024 | 0.009 | 0.538 |
| F60, Personality | -0.013 | -0.030 | 0.004 | 0.357 |
| F80, Speech and language | 0.016 | -0.001 | 0.033 | 0.194 |
| F81, Scholastic skills | 0.006 | -0.010 | 0.022 | 0.618 |
| Autism | 0.026 | 0.010 | 0.042 | 0.009 |
| ADHD | 0.047 | 0.031 | 0.063 | <0.001 |
| F93, Childhood emotional | 0.001 | -0.015 | 0.017 | 0.922 |
| F98, Child or adolescent (other) | 0.008 | -0.008 | 0.023 | 0.518 |
| F99, Unspecified | 0.009 | -0.007 | 0.025 | 0.448 |

*Note:* ADHD = attention-deficit/hyperactivity disorder; CI = confidence interval. Models were adjusted for chronological age, sex, assessment centre, batch number, the first six genetic principal components and fasting time. Cells highlighted in grey and blue correspond to statistically significant associations (nominally and after multiple testing corrections, respectively). *P*-values shown are corrected for multiple testing using the Benjamini–Hochberg procedure. $N = 219{,}494$.

**Table S10.** Ancestry-specific associations between MileAge delta and polygenic scores for individual disorders

| Polygenic score | Population | *β* | 95% CI | | *p* |
|---|---|---|---|---|---|
| F10, Alcohol use | AFR | 0.071 | -0.071 | 0.213 | 0.817 |
| | AMR | -0.842 | -1.635 | -0.049 | 0.422 |
| | CSA | 0.000 | -0.132 | 0.133 | 0.995 |
| | EAS | -0.246 | -0.537 | 0.046 | 0.637 |
| | EUR | -0.008 | -0.026 | 0.009 | 0.488 |
| | MID | 0.906 | -0.173 | 1.985 | 0.520 |
| F11, Opioid use | AFR | 0.071 | -0.086 | 0.229 | 0.817 |
| | AMR | 0.157 | -0.526 | 0.840 | 0.777 |
| | CSA | -0.064 | -0.196 | 0.068 | 0.608 |
| | EAS | -0.017 | -0.306 | 0.271 | 0.963 |
| | EUR | -0.009 | -0.026 | 0.009 | 0.488 |
| | MID | -0.047 | -1.112 | 1.017 | 0.935 |
| F12, Cannabis use | AFR | 0.049 | -0.092 | 0.189 | 0.817 |
| | AMR | 0.068 | -0.556 | 0.692 | 0.907 |
| | CSA | 0.117 | -0.011 | 0.245 | 0.557 |
| | EAS | -0.054 | -0.338 | 0.230 | 0.957 |
| | EUR | -0.002 | -0.019 | 0.015 | 0.943 |
| | MID | -0.282 | -1.236 | 0.672 | 0.854 |
| F13, Sedative use | AFR | -0.105 | -0.242 | 0.033 | 0.817 |
| | AMR | 0.327 | -0.350 | 1.003 | 0.610 |
| | CSA | -0.065 | -0.188 | 0.058 | 0.608 |
| | EAS | 0.115 | -0.155 | 0.385 | 0.957 |
| | EUR | 0.000 | -0.016 | 0.016 | 0.992 |
| | MID | -0.588 | -1.693 | 0.517 | 0.753 |
| F15, Stimulant use | AFR | -0.018 | -0.154 | 0.118 | 0.900 |
| | AMR | -0.266 | -1.366 | 0.834 | 0.777 |
| | CSA | -0.102 | -0.236 | 0.032 | 0.557 |
| | EAS | -0.054 | -0.353 | 0.245 | 0.957 |
| | EUR | 0.001 | -0.016 | 0.019 | 0.978 |
| | MID | 0.681 | -0.237 | 1.600 | 0.664 |
| F17, Tobacco use | AFR | -0.050 | -0.184 | 0.084 | 0.817 |
| | AMR | -0.522 | -1.112 | 0.068 | 0.422 |
| | CSA | -0.009 | -0.138 | 0.121 | 0.995 |
| | EAS | -0.014 | -0.309 | 0.281 | 0.963 |
| | EUR | -0.034 | -0.051 | -0.017 | <0.001 |
| | MID | -0.147 | -0.996 | 0.702 | 0.872 |
| F19, Multi-substance use | AFR | 0.029 | -0.107 | 0.164 | 0.887 |
| | AMR | 0.284 | -0.635 | 1.203 | 0.746 |
| | CSA | 0.035 | -0.098 | 0.167 | 0.898 |
| | EAS | -0.245 | -0.539 | 0.050 | 0.637 |
| | EUR | -0.001 | -0.018 | 0.017 | 0.980 |
| | MID | -0.261 | -1.145 | 0.623 | 0.854 |
| Schizophrenia | AFR | -0.027 | -0.182 | 0.127 | 0.894 |
| | AMR | 0.466 | -0.095 | 1.027 | 0.422 |
| | CSA | 0.029 | -0.108 | 0.166 | 0.928 |
| | EAS | 0.074 | -0.221 | 0.369 | 0.957 |
| | EUR | -0.012 | -0.028 | 0.004 | 0.292 |
| | MID | 0.059 | -1.103 | 1.220 | 0.935 |
| F22, Delusional | AFR | 0.056 | -0.095 | 0.206 | 0.817 |
| | AMR | 0.242 | -0.433 | 0.918 | 0.739 |
| | CSA | -0.004 | -0.146 | 0.139 | 0.995 |
| | EAS | 0.261 | -0.030 | 0.553 | 0.637 |
| | EUR | -0.023 | -0.038 | -0.007 | 0.020 |
| | MID | 0.571 | -0.362 | 1.505 | 0.743 |
| F23, Acute psychosis | AFR | 0.005 | -0.138 | 0.147 | 0.974 |
| | AMR | -0.635 | -1.300 | 0.030 | 0.422 |
| | CSA | -0.009 | -0.140 | 0.123 | 0.995 |
| | EAS | 0.133 | -0.160 | 0.426 | 0.957 |
| | EUR | -0.042 | -0.059 | -0.025 | <0.001 |
| | MID | -0.222 | -1.200 | 0.755 | 0.854 |
| F25, Schizoaffective | AFR | 0.010 | -0.129 | 0.149 | 0.956 |
| | AMR | -0.431 | -1.110 | 0.249 | 0.514 |
| | CSA | -0.020 | -0.145 | 0.105 | 0.928 |
| | EAS | 0.124 | -0.164 | 0.412 | 0.957 |

| | | | | | |
|---|---|---|---|---|---|
| | EUR | -0.024 | -0.040 | -0.008 | 0.020 |
| | MID | 0.258 | -0.875 | 1.391 | 0.854 |
| F29, Psychosis (unspecific) | AFR | 0.009 | -0.137 | 0.155 | 0.956 |
| | AMR | 0.254 | -0.632 | 1.141 | 0.746 |
| | CSA | -0.092 | -0.218 | 0.034 | 0.557 |
| | EAS | 0.055 | -0.246 | 0.356 | 0.957 |
| | EUR | -0.028 | -0.045 | -0.011 | 0.006 |
| | MID | 1.058 | -0.111 | 2.228 | 0.465 |
| F30, Mania | AFR | 0.053 | -0.095 | 0.201 | 0.817 |
| | AMR | 0.172 | -0.448 | 0.792 | 0.746 |
| | CSA | 0.030 | -0.099 | 0.160 | 0.920 |
| | EAS | 0.050 | -0.231 | 0.331 | 0.957 |
| | EUR | -0.006 | -0.023 | 0.012 | 0.682 |
| | MID | -0.305 | -1.280 | 0.670 | 0.854 |
| Bipolar | AFR | 0.060 | -0.102 | 0.223 | 0.817 |
| | AMR | 0.510 | -0.198 | 1.218 | 0.446 |
| | CSA | -0.003 | -0.130 | 0.124 | 0.995 |
| | EAS | 0.310 | 0.024 | 0.597 | 0.637 |
| | EUR | -0.005 | -0.021 | 0.010 | 0.682 |
| | MID | 0.343 | -0.764 | 1.450 | 0.854 |
| F32, Depressive episode | AFR | -0.045 | -0.190 | 0.099 | 0.817 |
| | AMR | 0.464 | -0.144 | 1.073 | 0.446 |
| | CSA | 0.097 | -0.039 | 0.232 | 0.557 |
| | EAS | -0.169 | -0.457 | 0.120 | 0.957 |
| | EUR | 0.011 | -0.006 | 0.027 | 0.392 |
| | MID | 0.150 | -0.891 | 1.192 | 0.879 |
| F33, Recurrent depression | AFR | -0.002 | -0.146 | 0.141 | 0.975 |
| | AMR | 0.581 | -0.099 | 1.260 | 0.422 |
| | CSA | 0.035 | -0.096 | 0.166 | 0.898 |
| | EAS | 0.040 | -0.267 | 0.348 | 0.957 |
| | EUR | 0.004 | -0.013 | 0.020 | 0.829 |
| | MID | 0.279 | -0.542 | 1.100 | 0.854 |
| Depression | AFR | 0.025 | -0.127 | 0.176 | 0.894 |
| | AMR | 0.503 | -0.176 | 1.183 | 0.446 |
| | CSA | 0.081 | -0.058 | 0.220 | 0.557 |
| | EAS | -0.069 | -0.355 | 0.216 | 0.957 |
| | EUR | 0.019 | 0.003 | 0.034 | 0.078 |
| | MID | 0.891 | -0.073 | 1.854 | 0.465 |
| F34, Persistent mood | AFR | -0.083 | -0.222 | 0.055 | 0.817 |
| | AMR | 0.308 | -0.328 | 0.944 | 0.610 |
| | CSA | -0.094 | -0.223 | 0.035 | 0.557 |
| | EAS | 0.012 | -0.259 | 0.283 | 0.963 |
| | EUR | -0.010 | -0.027 | 0.007 | 0.417 |
| | MID | -0.108 | -0.988 | 0.772 | 0.879 |
| F39, Mood (unspecific) | AFR | 0.032 | -0.116 | 0.181 | 0.887 |
| | AMR | 0.027 | -0.548 | 0.603 | 0.925 |
| | CSA | 0.066 | -0.064 | 0.197 | 0.608 |
| | EAS | 0.243 | -0.038 | 0.523 | 0.637 |
| | EUR | 0.010 | -0.007 | 0.027 | 0.417 |
| | MID | -0.463 | -1.587 | 0.661 | 0.854 |
| F40, Phobic | AFR | 0.082 | -0.059 | 0.224 | 0.817 |
| | AMR | 0.493 | -0.249 | 1.235 | 0.506 |
| | CSA | 0.082 | -0.050 | 0.214 | 0.557 |
| | EAS | -0.049 | -0.355 | 0.257 | 0.957 |
| | EUR | -0.015 | -0.033 | 0.002 | 0.239 |
| | MID | -0.048 | -1.235 | 1.138 | 0.935 |
| F41, Anxiety | AFR | 0.018 | -0.123 | 0.159 | 0.900 |
| | AMR | 0.601 | -0.102 | 1.303 | 0.422 |
| | CSA | 0.086 | -0.049 | 0.220 | 0.557 |
| | EAS | -0.046 | -0.337 | 0.246 | 0.957 |
| | EUR | -0.011 | -0.027 | 0.005 | 0.346 |
| | MID | -0.203 | -1.349 | 0.943 | 0.872 |
| F42, Obsessive-compulsive | AFR | -0.042 | -0.179 | 0.095 | 0.817 |
| | AMR | -0.158 | -0.696 | 0.381 | 0.746 |
| | CSA | -0.076 | -0.202 | 0.050 | 0.557 |
| | EAS | -0.035 | -0.324 | 0.254 | 0.957 |

| | | | | | |
|---|---|---|---|---|---|
| | EUR | -0.035 | -0.051 | -0.018 | <0.001 |
| | MID | -0.299 | -1.207 | 0.609 | 0.854 |
| F43, Stress-related | AFR | 0.118 | -0.032 | 0.267 | 0.817 |
| | AMR | -0.252 | -0.851 | 0.348 | 0.687 |
| | CSA | -0.078 | -0.204 | 0.048 | 0.557 |
| | EAS | -0.168 | -0.477 | 0.140 | 0.957 |
| | EUR | -0.010 | -0.027 | 0.007 | 0.435 |
| | MID | -0.547 | -1.512 | 0.417 | 0.743 |
| F44, Dissociative | AFR | 0.062 | -0.085 | 0.209 | 0.817 |
| | AMR | -0.435 | -1.027 | 0.158 | 0.446 |
| | CSA | 0.082 | -0.050 | 0.214 | 0.557 |
| | EAS | -0.244 | -0.558 | 0.071 | 0.683 |
| | EUR | 0.003 | -0.013 | 0.020 | 0.829 |
| | MID | -0.215 | -1.213 | 0.784 | 0.854 |
| F45, Somatoform | AFR | 0.062 | -0.082 | 0.206 | 0.817 |
| | AMR | 0.033 | -0.595 | 0.660 | 0.925 |
| | CSA | 0.004 | -0.127 | 0.135 | 0.995 |
| | EAS | 0.161 | -0.153 | 0.474 | 0.957 |
| | EUR | -0.015 | -0.032 | 0.002 | 0.217 |
| | MID | 0.237 | -0.650 | 1.125 | 0.854 |
| F48, Other neurotic | AFR | 0.098 | -0.039 | 0.235 | 0.817 |
| | AMR | 0.777 | 0.122 | 1.432 | 0.422 |
| | CSA | 0.022 | -0.101 | 0.145 | 0.928 |
| | EAS | 0.254 | -0.018 | 0.527 | 0.637 |
| | EUR | -0.014 | -0.030 | 0.003 | 0.242 |
| | MID | -0.199 | -1.101 | 0.703 | 0.854 |
| F50, Eating | AFR | 0.045 | -0.097 | 0.187 | 0.817 |
| | AMR | 0.291 | -0.317 | 0.900 | 0.610 |
| | CSA | -0.062 | -0.192 | 0.067 | 0.608 |
| | EAS | 0.099 | -0.183 | 0.380 | 0.957 |
| | EUR | 0.001 | -0.015 | 0.018 | 0.978 |
| | MID | 1.052 | -0.027 | 2.131 | 0.465 |
| Anorexia | AFR | -0.095 | -0.261 | 0.071 | 0.817 |
| | AMR | 0.239 | -0.565 | 1.043 | 0.746 |
| | CSA | -0.053 | -0.190 | 0.084 | 0.719 |
| | EAS | -0.089 | -0.380 | 0.203 | 0.957 |
| | EUR | -0.022 | -0.037 | -0.007 | 0.020 |
| | MID | -0.165 | -1.483 | 1.152 | 0.879 |
| F51, Sleep | AFR | 0.042 | -0.096 | 0.180 | 0.817 |
| | AMR | 0.566 | -0.103 | 1.235 | 0.422 |
| | CSA | 0.050 | -0.077 | 0.176 | 0.719 |
| | EAS | -0.106 | -0.394 | 0.183 | 0.957 |
| | EUR | -0.009 | -0.026 | 0.007 | 0.435 |
| | MID | 1.126 | 0.262 | 1.989 | 0.409 |
| F60, Personality | AFR | 0.029 | -0.110 | 0.167 | 0.887 |
| | AMR | 0.843 | 0.054 | 1.632 | 0.422 |
| | CSA | 0.078 | -0.056 | 0.212 | 0.557 |
| | EAS | -0.042 | -0.329 | 0.245 | 0.957 |
| | EUR | -0.017 | -0.034 | 0.001 | 0.182 |
| | MID | 0.663 | -0.396 | 1.721 | 0.743 |
| F80, Speech and language | AFR | -0.045 | -0.191 | 0.102 | 0.817 |
| | AMR | -0.382 | -1.035 | 0.272 | 0.514 |
| | CSA | -0.089 | -0.220 | 0.041 | 0.557 |
| | EAS | -0.136 | -0.415 | 0.142 | 0.957 |
| | EUR | 0.019 | 0.002 | 0.036 | 0.093 |
| | MID | -0.614 | -1.664 | 0.437 | 0.743 |
| F81, Scholastic skills | AFR | -0.030 | -0.182 | 0.121 | 0.887 |
| | AMR | 0.241 | -0.405 | 0.887 | 0.739 |
| | CSA | 0.122 | -0.003 | 0.247 | 0.557 |
| | EAS | 0.038 | -0.259 | 0.336 | 0.957 |
| | EUR | 0.004 | -0.013 | 0.020 | 0.829 |
| | MID | -1.301 | -2.409 | -0.193 | 0.409 |
| Autism | AFR | -0.106 | -0.251 | 0.040 | 0.817 |
| | AMR | -0.064 | -0.724 | 0.595 | 0.907 |
| | CSA | -0.001 | -0.129 | 0.127 | 0.995 |
| | EAS | -0.089 | -0.379 | 0.201 | 0.957 |

| | | | | | |
|---|---|---|---|---|---|
| | EUR | 0.028 | 0.012 | 0.044 | 0.006 |
| | MID | 0.658 | -0.342 | 1.658 | 0.743 |
| ADHD | AFR | 0.211 | 0.076 | 0.346 | 0.079 |
| | AMR | 0.060 | -0.605 | 0.725 | 0.907 |
| | CSA | 0.109 | -0.022 | 0.240 | 0.557 |
| | EAS | -0.073 | -0.378 | 0.232 | 0.957 |
| | EUR | 0.042 | 0.026 | 0.059 | <0.001 |
| | MID | 0.389 | -0.635 | 1.413 | 0.854 |
| F93, Childhood emotional | AFR | 0.050 | -0.098 | 0.197 | 0.817 |
| | AMR | 0.355 | -0.249 | 0.959 | 0.514 |
| | CSA | 0.116 | -0.012 | 0.243 | 0.557 |
| | EAS | -0.011 | -0.285 | 0.263 | 0.963 |
| | EUR | -0.001 | -0.017 | 0.016 | 0.980 |
| | MID | -1.025 | -2.007 | -0.043 | 0.465 |
| F98, Child or adolescent (other) | AFR | -0.080 | -0.214 | 0.053 | 0.817 |
| | AMR | 0.461 | -0.282 | 1.203 | 0.514 |
| | CSA | -0.088 | -0.214 | 0.037 | 0.557 |
| | EAS | -0.007 | -0.305 | 0.291 | 0.965 |
| | EUR | 0.012 | -0.004 | 0.028 | 0.292 |
| | MID | 0.251 | -0.891 | 1.393 | 0.854 |
| F99, Unspecified | AFR | -0.054 | -0.195 | 0.086 | 0.817 |
| | AMR | -0.137 | -0.834 | 0.561 | 0.809 |
| | CSA | 0.024 | -0.108 | 0.157 | 0.928 |
| | EAS | 0.029 | -0.236 | 0.295 | 0.957 |
| | EUR | 0.009 | -0.008 | 0.025 | 0.456 |
| | MID | 0.658 | -0.614 | 1.930 | 0.753 |

*Note:* AFR = African; AMR = Admixed American; EAS = East Asian; EUR = European; CSA = Central and South Asian; MID = Middle Eastern. Models were adjusted for chronological age, sex, assessment centre, batch number (except for MID), the first six genetic principal components and fasting time. Cells highlighted in grey and blue correspond to statistically significant associations (nominally and after multiple testing corrections, respectively). *P*-values shown are corrected for multiple testing using the Benjamini–Hochberg procedure (separately within each population). $N = 3426$ (AFR); $N = 278$ (AMR); $N = 3916$ (CSA); $N = 1012$ (EAS); $N = 210{,}755$ (EUR); $N = 94$ (MID).

#### 12. MileAge delta and individual two-digit ICD-10 codes; excluding self-report

**Table S11.** MileAge delta and mental/behavioural disorders; excluding self-report diagnoses

| | | Model 1 (adj. age and sex) | | | | Model 2 (full adjustment) | | | |
|---|---|---|---|---|---|---|---|---|---|
| ICD-10 | *N* | *β* | 95% CI | | *p* | *β* | 95% CI | | *p* |
| None | 186688 | Reference | | | | Reference | | | |
| F10, Alcohol use | 1303 | 0.705 | 0.500 | 0.910 | <0.001 | 0.603 | 0.397 | 0.808 | <0.001 |
| F11, Opioid use | 46 | 1.113 | 0.028 | 2.199 | 0.157 | 0.907 | -0.178 | 1.992 | 0.285 |
| F12, Cannabis use | 46 | -0.914 | -1.999 | 0.172 | 0.285 | -1.108 | -2.193 | -0.023 | 0.157 |
| F13, Sedative use | 68 | -0.394 | -1.287 | 0.498 | 0.571 | -0.479 | -1.371 | 0.413 | 0.537 |
| F14, Cocaine use | 10 | 0.965 | -1.363 | 3.293 | 0.595 | 0.710 | -1.616 | 3.036 | 0.707 |
| F15, Stimulant use | 14 | 0.942 | -1.026 | 2.909 | 0.569 | 0.739 | -1.227 | 2.705 | 0.615 |
| F16, Hallucinogen use | 12 | 3.758 | 1.633 | 5.883 | 0.006 | 3.630 | 1.507 | 5.753 | 0.008 |
| F17, Tobacco use | 9715 | 0.409 | 0.332 | 0.485 | <0.001 | 0.373 | 0.296 | 0.450 | <0.001 |
| F19, Multi-substance use | 61 | 0.598 | -0.344 | 1.541 | 0.497 | 0.440 | -0.502 | 1.382 | 0.569 |
| F20, Schizophrenia | 150 | 0.453 | -0.148 | 1.055 | 0.379 | 0.236 | -0.366 | 0.837 | 0.615 |
| F22, Delusional | 88 | 0.956 | 0.171 | 1.741 | 0.082 | 0.804 | 0.020 | 1.589 | 0.157 |
| F23, Acute psychosis | 66 | 0.044 | -0.862 | 0.950 | 0.956 | -0.069 | -0.974 | 0.837 | 0.937 |
| F25, Schizoaffective | 85 | 0.905 | 0.106 | 1.703 | 0.113 | 0.729 | -0.069 | 1.527 | 0.236 |
| F29, Psychosis (unspecific) | 83 | 0.541 | -0.267 | 1.349 | 0.461 | 0.366 | -0.442 | 1.174 | 0.569 |
| F30, Mania | 124 | 0.494 | -0.167 | 1.155 | 0.379 | 0.408 | -0.252 | 1.069 | 0.497 |
| F31, Bipolar | 162 | 0.635 | 0.057 | 1.214 | 0.123 | 0.535 | -0.044 | 1.113 | 0.233 |
| F32, Depressive episode | 5425 | 0.288 | 0.186 | 0.389 | <0.001 | 0.243 | 0.141 | 0.345 | <0.001 |
| F33, Recurrent depression | 1211 | 0.327 | 0.115 | 0.539 | 0.017 | 0.268 | 0.056 | 0.480 | 0.070 |
| F34, Persistent mood | 306 | 0.247 | -0.174 | 0.668 | 0.501 | 0.190 | -0.231 | 0.611 | 0.569 |
| F38, Mood (other) | 36 | 0.300 | -0.927 | 1.527 | 0.748 | 0.237 | -0.989 | 1.462 | 0.814 |
| F39, Mood (unspecific) | 282 | 0.201 | -0.238 | 0.640 | 0.569 | 0.171 | -0.268 | 0.609 | 0.615 |
| F40, Phobic | 717 | 0.151 | -0.125 | 0.426 | 0.531 | 0.119 | -0.156 | 0.395 | 0.575 |
| F41, Anxiety | 4772 | 0.206 | 0.098 | 0.314 | 0.002 | 0.170 | 0.062 | 0.278 | 0.017 |
| F42, Obsessive-compulsive | 214 | -0.698 | -1.201 | -0.194 | 0.037 | -0.776 | -1.279 | -0.273 | 0.017 |
| F43, Stress-related | 4094 | 0.193 | 0.077 | 0.310 | 0.010 | 0.167 | 0.051 | 0.283 | 0.030 |
| F44, Dissociative | 79 | 0.374 | -0.454 | 1.203 | 0.569 | 0.317 | -0.510 | 1.145 | 0.615 |
| F45, Somatoform | 843 | 0.021 | -0.233 | 0.276 | 0.937 | 0.017 | -0.237 | 0.271 | 0.937 |
| F48, Other neurotic | 108 | -0.318 | -1.026 | 0.391 | 0.569 | -0.348 | -1.056 | 0.360 | 0.569 |
| F50, Eating | 182 | -0.245 | -0.792 | 0.301 | 0.569 | -0.257 | -0.803 | 0.289 | 0.569 |
| F51, Sleep | 153 | 0.367 | -0.228 | 0.963 | 0.497 | 0.362 | -0.232 | 0.957 | 0.498 |
| F52, Sexual dysfunction | 632 | -0.008 | -0.301 | 0.286 | 0.971 | -0.012 | -0.305 | 0.281 | 0.959 |
| F53, Puerperal psychosis | 268 | -0.393 | -0.843 | 0.058 | 0.272 | -0.383 | -0.833 | 0.067 | 0.285 |
| F60, Personality | 180 | 0.805 | 0.256 | 1.354 | 0.026 | 0.667 | 0.118 | 1.216 | 0.082 |
| F63, Impulse | 16 | 1.142 | -0.698 | 2.982 | 0.497 | 1.083 | -0.755 | 2.922 | 0.501 |
| F66, Sexual development | 155 | -0.333 | -0.924 | 0.259 | 0.524 | -0.351 | -0.942 | 0.240 | 0.501 |
| F69, Personality (unspecific) | 64 | -0.003 | -0.923 | 0.918 | 0.996 | -0.089 | -1.009 | 0.830 | 0.932 |
| F80, Speech and language | 31 | 0.441 | -0.881 | 1.763 | 0.670 | 0.375 | -0.946 | 1.696 | 0.712 |
| F81, Scholastic skills | 60 | 0.355 | -0.596 | 1.305 | 0.615 | 0.236 | -0.714 | 1.186 | 0.748 |
| F84, Pervasive developmental | 18 | 0.497 | -1.238 | 2.232 | 0.712 | 0.357 | -1.376 | 2.091 | 0.802 |
| F90, ADHD | 10 | -0.324 | -2.652 | 2.003 | 0.883 | -0.373 | -2.699 | 1.952 | 0.858 |
| F91, Conduct | 43 | 1.314 | 0.192 | 2.437 | 0.098 | 1.238 | 0.117 | 2.360 | 0.123 |
| F93, Childhood emotional | 11 | 0.650 | -1.570 | 2.869 | 0.712 | 0.594 | -1.624 | 2.811 | 0.729 |
| F95, Tic | 11 | 1.550 | -0.669 | 3.770 | 0.431 | 1.544 | -0.673 | 3.761 | 0.431 |
| F98, Child or adolescent (other) | 45 | -0.580 | -1.678 | 0.517 | 0.540 | -0.612 | -1.709 | 0.484 | 0.524 |
| F99, Unspecified | 86 | 0.085 | -0.708 | 0.879 | 0.925 | -0.059 | -0.852 | 0.735 | 0.937 |

*Note:* ICD-10 = International Classification of Diseases, 10th Revision; CI = confidence interval. Model 1–adjusted for chronological age and sex; Model 2–adjusted for chronological age, sex, ethnicity, cohabitation with spouse/partner, highest educational/professional qualification, annual gross household income, Townsend deprivation index and fasting time. For diagnoses printed in grey, there were no differences in sample size compared to the primary analysis. Cells highlighted in grey and blue correspond to statistically significant associations (nominally and after multiple testing corrections, respectively). *P*-values shown are corrected for multiple testing using the Benjamini–Hochberg procedure.

#### 13. MileAge delta and groups of two-digit ICD-10 codes; primary care linkage

**Table S12.** MileAge delta and mental/behavioural disorders; primary care linkage

| | | Model 1 (adj. age and sex) | | | | Model 2 (full adjustment) | | | |
|---|---|---|---|---|---|---|---|---|---|
| ICD-10 | | *β* | 95% CI | | *p* | *β* | 95% CI | | *p* |
| None | 75576 | Reference | | | | Reference | | | |
| F10-99, Any disorder | 27147 | 0.226 | 0.174 | 0.279 | <0.001 | 0.197 | 0.144 | 0.250 | <0.001 |
| F10-19, Substance use | 10265 | 0.379 | 0.302 | 0.457 | <0.001 | 0.352 | 0.274 | 0.431 | <0.001 |
| F20-29, Psychosis | 319 | 0.716 | 0.304 | 1.128 | 0.002 | 0.597 | 0.183 | 1.010 | 0.010 |
| F30-39, Affective | 11834 | 0.241 | 0.168 | 0.314 | <0.001 | 0.205 | 0.131 | 0.279 | <0.001 |
| F40-48, Neurotic | 10900 | 0.136 | 0.060 | 0.211 | 0.001 | 0.111 | 0.035 | 0.187 | 0.010 |
| F50-59, Behavioural syndromes | 1399 | -0.243 | -0.442 | -0.045 | 0.030 | -0.245 | -0.443 | -0.046 | 0.030 |
| F60-69, Personality & behaviour | 368 | 0.154 | -0.230 | 0.538 | 0.500 | 0.109 | -0.275 | 0.493 | 0.636 |
| F70-79, Mental retardation | 11 | 1.559 | -0.656 | 3.773 | 0.231 | 1.392 | -0.822 | 3.607 | 0.266 |
| F80-89, Developmental | 90 | 0.593 | -0.182 | 1.368 | 0.210 | 0.506 | -0.269 | 1.282 | 0.260 |
| F90-98, Child & adolescent | 118 | 0.542 | -0.135 | 1.218 | 0.197 | 0.498 | -0.178 | 1.174 | 0.218 |
| F99, Unspecified | 56 | 0.180 | -0.802 | 1.161 | 0.754 | 0.085 | -0.897 | 1.066 | 0.865 |

*Note:* ICD-10 = International Classification of Diseases, 10th Revision; CI = confidence interval. Model 1–adjusted for chronological age and sex; Model 2–adjusted for chronological age, sex, ethnicity, cohabitation with spouse/partner, highest educational/professional qualification, annual gross household income, Townsend deprivation index and fasting time. Cells highlighted in grey and blue correspond to statistically significant associations (nominally and after multiple testing corrections, respectively). *P*-values shown are corrected for multiple testing using the Benjamini–Hochberg procedure.

### 14. MileAge delta and individual two-digit ICD-10 codes; primary care linkage

**Table S13.** MileAge delta and mental/behavioural disorders; primary care linkage

| | | Model 1 (adj. age and sex) | | | | Model 2 (full adjustment) | | | |
|---|---|---|---|---|---|---|---|---|---|
| ICD-10 | *N* | *β* | 95% CI | | *p* | *β* | 95% CI | | *p* |
| None | 75576 | Reference | | | | Reference | | | |
| F10, Alcohol use | 839 | 0.612 | 0.357 | 0.868 | <0.001 | 0.543 | 0.286 | 0.800 | <0.001 |
| F11, Opioid use | 29 | 1.025 | -0.339 | 2.389 | 0.417 | 0.885 | -0.479 | 2.250 | 0.461 |
| F12, Cannabis use | 30 | -0.826 | -2.168 | 0.515 | 0.489 | -0.913 | -2.256 | 0.429 | 0.436 |
| F13, Sedative use | 66 | -0.438 | -1.342 | 0.467 | 0.602 | -0.499 | -1.403 | 0.406 | 0.519 |
| F14, Cocaine use | | *N* insufficient | | | | | | | |
| F15, Stimulant use | 11 | 0.982 | -1.232 | 3.197 | 0.642 | 0.795 | -1.419 | 3.009 | 0.690 |
| F16, Hallucinogen use | | *N* insufficient | | | | | | | |
| F17, Tobacco use | 9556 | 0.365 | 0.285 | 0.445 | <0.001 | 0.342 | 0.261 | 0.423 | <0.001 |
| F19, Multi-substance use | 59 | 0.676 | -0.281 | 1.633 | 0.436 | 0.561 | -0.396 | 1.518 | 0.513 |
| F20, Schizophrenia | 182 | 1.020 | 0.474 | 1.565 | 0.003 | 0.866 | 0.319 | 1.413 | 0.021 |
| F22, Delusional | 66 | 0.909 | 0.005 | 1.814 | 0.168 | 0.797 | -0.108 | 1.702 | 0.278 |
| F23, Acute psychosis | 43 | -0.307 | -1.428 | 0.813 | 0.758 | -0.376 | -1.496 | 0.744 | 0.708 |
| F25, Schizoaffective | 52 | 0.806 | -0.213 | 1.825 | 0.385 | 0.700 | -0.319 | 1.719 | 0.436 |
| F29, Psychosis (unspecific) | 66 | 0.293 | -0.612 | 1.198 | 0.717 | 0.157 | -0.748 | 1.062 | 0.833 |
| F30, Mania | 99 | 0.374 | -0.364 | 1.113 | 0.574 | 0.321 | -0.418 | 1.059 | 0.642 |
| F31, Bipolar | 362 | 0.168 | -0.219 | 0.555 | 0.642 | 0.099 | -0.288 | 0.487 | 0.768 |
| F32, Depressive episode | 10933 | 0.253 | 0.178 | 0.329 | <0.001 | 0.217 | 0.140 | 0.293 | <0.001 |
| F33, Recurrent depression | 1072 | 0.286 | 0.060 | 0.512 | 0.075 | 0.240 | 0.014 | 0.466 | 0.148 |
| F34, Persistent mood | 292 | 0.175 | -0.256 | 0.605 | 0.676 | 0.124 | -0.307 | 0.555 | 0.756 |
| F38, Mood (other) | 36 | 0.273 | -0.952 | 1.497 | 0.802 | 0.214 | -1.010 | 1.438 | 0.833 |
| F39, Mood (unspecific) | 273 | 0.179 | -0.267 | 0.624 | 0.676 | 0.152 | -0.293 | 0.598 | 0.708 |
| F40, Phobic | 666 | 0.034 | -0.253 | 0.320 | 0.880 | 0.009 | -0.277 | 0.295 | 0.971 |
| F41, Anxiety | 6076 | 0.135 | 0.037 | 0.234 | 0.049 | 0.106 | 0.007 | 0.204 | 0.148 |
| F42, Obsessive-compulsive | 218 | -0.600 | -1.098 | -0.102 | 0.098 | -0.666 | -1.164 | -0.167 | 0.058 |
| F43, Stress-related | 4336 | 0.168 | 0.053 | 0.283 | 0.034 | 0.150 | 0.035 | 0.265 | 0.067 |
| F44, Dissociative | 57 | 0.236 | -0.737 | 1.209 | 0.780 | 0.175 | -0.798 | 1.148 | 0.833 |
| F45, Somatoform | 770 | -0.040 | -0.306 | 0.226 | 0.842 | -0.046 | -0.312 | 0.220 | 0.833 |
| F48, Other neurotic | 266 | -0.175 | -0.626 | 0.276 | 0.688 | -0.214 | -0.665 | 0.237 | 0.607 |
| F50, Eating | 266 | -0.464 | -0.916 | -0.012 | 0.159 | -0.479 | -0.931 | -0.027 | 0.148 |
| F51, Sleep | 153 | 0.331 | -0.263 | 0.926 | 0.519 | 0.325 | -0.269 | 0.919 | 0.519 |
| F52, Sexual dysfunction | 569 | -0.112 | -0.421 | 0.197 | 0.690 | -0.113 | -0.422 | 0.195 | 0.690 |
| F53, Puerperal psychosis | 430 | -0.530 | -0.886 | -0.174 | 0.034 | -0.519 | -0.875 | -0.163 | 0.034 |
| F60, Personality | 126 | 0.766 | 0.111 | 1.421 | 0.104 | 0.683 | 0.027 | 1.339 | 0.154 |
| F63, Impulse | 16 | 1.137 | -0.699 | 2.974 | 0.489 | 1.083 | -0.752 | 2.918 | 0.513 |
| F66, Sexual development | 153 | -0.396 | -0.990 | 0.199 | 0.446 | -0.413 | -1.007 | 0.181 | 0.436 |
| F69, Personality (unspecific) | 59 | -0.050 | -1.007 | 0.906 | 0.951 | -0.100 | -1.056 | 0.856 | 0.890 |
| F80, Speech and language | 25 | 0.016 | -1.453 | 1.485 | 0.983 | -0.027 | -1.496 | 1.441 | 0.982 |
| F81, Scholastic skills | 43 | 0.857 | -0.264 | 1.977 | 0.412 | 0.781 | -0.340 | 1.903 | 0.436 |
| F84, Pervasive developmental | 17 | 0.661 | -1.120 | 2.443 | 0.690 | 0.521 | -1.260 | 2.302 | 0.756 |
| F90, ADHD | 10 | -0.341 | -2.664 | 1.981 | 0.842 | -0.382 | -2.703 | 1.939 | 0.834 |
| F91, Conduct | 42 | 1.351 | 0.217 | 2.484 | 0.099 | 1.291 | 0.158 | 2.424 | 0.116 |
| F93, Childhood emotional | 11 | 0.625 | -1.590 | 2.839 | 0.756 | 0.577 | -1.636 | 2.790 | 0.768 |
| F95, Tic | 11 | 1.548 | -0.666 | 3.763 | 0.436 | 1.536 | -0.677 | 3.749 | 0.436 |
| F98, Child or adolescent (other) | 45 | -0.602 | -1.697 | 0.493 | 0.519 | -0.630 | -1.725 | 0.464 | 0.518 |
| F99, Unspecified | 56 | 0.180 | -0.802 | 1.161 | 0.833 | 0.085 | -0.897 | 1.066 | 0.908 |

*Note:* ICD-10 = International Classification of Diseases, 10th Revision; CI = confidence interval. Model 1–adjusted for chronological age and sex; Model 2–adjusted for chronological age, sex, ethnicity, cohabitation with spouse/partner, highest educational/professional qualification, annual gross household income, Townsend deprivation index and fasting time. For diagnoses printed in grey, there were no differences in sample size compared to the primary analysis. Cells highlighted in grey and blue correspond to statistically significant associations (nominally and after multiple testing corrections, respectively). *P*-values shown are corrected for multiple testing using the Benjamini–Hochberg procedure.
